# A Surprising formula for the spread of Covid-19 Under Aggressive Management

**DOI:** 10.1101/2020.04.29.20084483

**Authors:** Ivan Cherednik

**Affiliations:** UNC at Chapel Hill

## Abstract

We propose an algebraic-type formula that describes with high accuracy the total number of detected infections for the Covid-19 pandemic in many countries. Our 2-phase formula can be used as a powerful forecasting tool. It is based on the author’s new theory of momentum management of epidemics; Bessel functions are employed. Its 3 parameters are the initial transmission rate, reflecting the viral fitness and “normal” frequency of contacts in the infected areas, and the intensity of prevention measures at phases 1, 2. Austria, Brazil, Germany, Japan, India, Israel, Italy, the Netherlands, Sweden, Switzerland, UK, and the USA are considered, including the second wave in the latter. The forecasting software is provided as a supplement (any groups of countries).

## 1. Our approach and main findings

A surprising *2-phase formula* describing very well the total number of detected infections of *Covid-19* during practically for the whole period of the spread in many countries is the main result. We model *momentum management of epidemics*, which can be defined as a system of measures aimed at reducing the epidemic spread by regulating their intensity on the basis of the latest total numbers of infections. The “hard measures”, such as detection and prompt isolation of infected people and closing the places where the spread is the most likely, combined with prompt response to the current numbers of cases, ensure a relatively fast saturation. Assuming that the saturation is reached, the second phase is when the hard measures are reduced. The accuracy of our formula is even more surprising for this phase, since many political, economic and other factors are involved. The precision of our 2-phase formula practically in *all* countries that reached phase 2 seems a real discovery. The recurrence of any kind due to abandoning the measures is not considered. The readers mostly interested in the final formulas and particular countries can go directly to Section 5; Section 6 is on the auto-forecasting for the USA.

### Focus on risk-management

We attribute the surprising similarity of the curves of the *total numbers of detected cases* in many countries to the uniformity of the measures employed and the ways people react to the data. These numbers mostly reflect *symptomatic cases* and are frequently underreported. However, as far as they influence the decisions of the authorities in charge and our own behavior, they proved to be sufficient, a strong confirmation of our “sociological” approach.

We see 3 basic types of management. The countries in the first group are determined to reach “double digit numbers” of new daily infections. The second group is when the reduction of hard measures begins upon the first signs of the stabilization of the daily numbers, which can be still very high; this is the switch to the (*AB*)–mode or (*B*)-mode in our model. The third group of countries is where “hard” measures are not employed systematically, which can be due to a variety of reasons, including insufficient capacities or political decisions. Some measures are always in place, for instance, self-isolation of those who think that they can be infected. Their prompt requests for medical help can work well in the countries with solid health-care systems, like Sweden. Also, travel restrictions, common everywhere, definitely reduce the spread.

The general theory remains essentially unchanged from its first posted variant (April 13). However only now *all* its main features are confirmed to occur practically. For instance, *both* Bessel-type solutions appeared necessary to model the spread in Brazil, Italy, Germany, Japan and some other countries. Also, the log (*t*)–saturation of solutions of type (*B*) is the key to “ending the epidemic” for phase 2.

### Power law of epidemics

The simplest equations for the spread of communicable diseases result in exponential growth of the number of infections, which is mostly applicable to the initial stages of epidemics. See e.g. [1, 2, 3, 4] here and below and [5] about some perspectives with *Covid-19*. We focus on the middle stages, where the growth is practically no greater than some power functions in time, which obviously requires a different approach and different equations.

Also, the *logistic* models of the spread, as well as their SIR, SID generalizations, assume that the number of infections is comparable with the whole population, which we do not impose. Major epidemics were not really of this kind during the last 100 years, mostly due to better disease control worldwide and general life improvements.

The reality now is the power-type growth of *total* number of infections *U*(*t*) after a possible short period of exponential growth, *Covid-19* included. Our approach is based on this assumption. We start with the equation *dU*(*t*)/*dt* = *c U*(*t*)/*t*, where the *basic transmission rate c* here is a combination of the transmission strength of the virus and the “normal” frequency of the contacts in the infected place.

Generally, *dU*(*t*)/*dt* is essentially is proportional to *U*(*t*) − *U*(*t* − *p*), where *p* is the period when the infected people spread the virus in the most intensive way. This difference is really proportional to *U*(*t*)/*t* if the growth of *U*(*t*) is basically linear. It models well the growth of *U*(*t*) even without this assumption. The reduction of our contacts with infected people over time seems the most likely reason for this.

Let us try to clarify this. If the total number of cases growths linearly, i.e. when the number of new daily infections is constant, this is not really a “trigger” for us psychologically. However, we react strongly if the growth is parabolic or so.

Also, if someone wants to “see” the current *trend* of the epidemic using *only* the total number of infections to date, then *U*(*t*)/*t* is the best way. The equation *dU*(*t*)/*dt* = *c U*(*t*)/*t* immediately gives that *U*(*t*) = *Ct*^*c*^ for some constant *C*, i.e. results in the power growth. Such “sociological approach” to the epidemic spread is quite natural in our work: the active managements of epidemics is clearly of sociological nature. This is different from other power laws for infectious diseases; compare e.g. with [3].

There can be other mechanisms for the “power law”, including the biological ones. Self-isolation of infected species is common … unless for *rabies* ; it can grow over time and can depend on the intensity of the spread. The replication processes for viruses is another reduction mechanism, but this is well beyond this article.

This is how we behave before (regardless of) any protection measures. After the management began, *U*(*t*) is not ∼ *t*^*c*^ anymore; Bessel functions begin to control the spread till the “technical saturation”, provided that the measures are “hard”. They can soften after the turning point, or what looks like a turning point. We can model this too; our formula describes the *second phase* with very high accuracy.

### Some details

The full theory is presented in [6]. It is strongly related to that from [7]; the differential equations are the same. We begin there with modeling the *news impact over time* The “news impact” is absolutely relevant here: the discussion of the epidemic by the authorities in charge and everywhere is one of the key factors influencing our understanding the situation and the reduction of our contacts. Similarly, the active prevention measures are analogous to *momentum trading*.

The coefficient *c* is one of the two main mathematical parameters of our theory. It can be seen as follows.

Before the prevention measures are implemented, approximately it reveals itself in the growth ∼ *t*^*c*^ of the total number of detected infections, where *t* is time. Mostly, *c* ≈ 2.2 ∸ 2.6 for *Covid-19*. This is after a short period of the exponential growth. Upon the active management, the growth quickly drops to *t*^*c*/2^, i.e. becomes essentially linear, which is part of our theory in [6]. Importantly our *u*(*t*) and *w*(*t*) behave as ∼ *t*^*c*^ for relatively small *t*, so Bessel functions can be used very well to model the initial stages, not only the middle period.

### Ending epidemics

The “power law” is only a starting point of our analysis. The main problem is of course to “add” here some mechanisms ending epidemics and those preventing their possible recurrence. These are major challenges, biologically, psychologically, sociologically and mathematically. One can expect this *megaproblem* to be well beyond the power law itself, but we demonstrate that mathematically there is a path based on Bessel functions. The power growth alone does not lead to any saturation, the Bessel functions are needed.

We are of course fully aware of the statistical nature of the problem, but the formula for the growth of the total number of infections we propose works almost with an accuracy of physics laws. This is very surprising for such stochastic processes as epidemics.

An important outcome of our modeling is that the measures of “hard type”, like detecting and isolating infected people and closing the places where the spread is almost inevitable, are the key for ending an epidemic. The most aggressive “hard way” is to employ such measures strictly proportionally to the current total numbers of detected infections, not to its derivative of any kind.

For instance, if the number of infections doubles during some period, then *testing-detection* must be increased 4–fold. Similarly, when we approach the saturation, where the number of new detected infections becomes close to zero, the “hard response” is to continue testing *linearly* for some time, i.e. performing the same number of tests every day in spite of almost zero number of new infections.

By contrast, the “soft response” is as follows. We act upon the current *average* number of infections, instead of the *absolute* numbers, i.e. the response is slower. For instance, testing and other measures will practically stop when there are no new “cases”. If “hard” measures are not used, then mathematically the epidemic will never reach the saturation. However coupling the “soft response” with “hard measures” does result in the saturation, though it may take longer.

### The *u, w*–formulas

Upon composing the corresponding differential equations and integrating <u>t</u>hem we obtain the following. The function 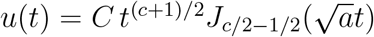 for the Bessel function *J*_*α*_ (*x*) of the first kind models the growth of the *total* number of infections, where *C* is the scaling parameter. Here and below time *t* is normalized *days/*10 for the number of days from the beginning of the period of the intense growth of the epidemic. Under “soft response” it is 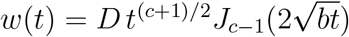 for the same *c* but different *D, b*. We will mostly discuss *u*(*t*), which work with high accuracy till the late stages. Both are ∼ *t*^*c*^ for small *t*. Thus both cover well the period of initial, essentially quadratic, growth of the number of detected infections. The parameters *a, b* give the *intensity* of “hard” measures correspondingly for *u*(*t*) and *w*(*t*).

For the *u*–function, *a*, the intensity (*A*)–measures, is 0.2 for the USA, UK, Italy and the Netherlands. It is 0.3 − 0.35 for Israel, Austria, Japan, Germany; 0.1 or smaller for Brazil, India, Sweden. The parameter *c*, the initial transmission rate, is 2.2 for the USA, 2.4 for Austria, the Netherlands, Sweden, UK, 2.6 for Italy, Germany, Japan. It jumps to around 5 in Brazil and India. This parameter can be captured at early stages; *a, b*(and *d* for phase 2) do depend on the management, but they are sufficiently uniform in quite a few countries.

### Limitations

Of course there can be other reasons for our *2-phase solution* to “serve” epidemics so well, not only due to the aggressive management. It is not impossible that there are connections with the replication process of viruses, but this we do not touch upon. As in any models, there are limitations, which we will address now.

First and foremost, the available infection numbers are for the *detected* cases, which are mostly *symptomatic*. However this is not of much concern to us. We understand managing epidemics from the viewpoint of the management. Infected people who are detected mostly have symptoms, but the change of the number of detected cases generally reflects well the “trend”, which is sufficient to properly adjust the intensity of the measures. So *de facto* focusing on symptomatic cases is basically sufficient for the management (and for our approach). No assumptions on asymptomatic cases are necessary to obtain our formulas. We of course understand that when the number of new *reported* infections drops to zero, there can be many non-detected asymptomatic cases, which can potentially lead to the recurrence of the epidemic. Such a saturation is only a *technical* end of the epidemic.

The second reservation concerns *newly emerged clusters of infections*, testing in new areas, and the countries where the spread is on the rise. The *u, w*–formulas can be used, but significant fluctuations can be expected. Also the usage of *both* Bessel-type solutions of our equations appeared necessary to address the “bulges” of the curves in quite a few countries. Anyway, the parameters and predictions must be constantly updated.

The third reservation is related to the management of the epidemic. Not all countries employ the protection measures in similar ways, but this is not a problem to us. The problem is if the intensity of the measures and the criteria are changed in some irregular ways. Diminishing the “hard” measures too early or even dropping them completely at later stages is of this kind; this is where *w*(*t*) comes into play.

Last but not the least is the *data quality*. Changing frequently the ways they are collected and the criteria makes such data useless for us. Though, if the number of detected infections is underreported in some regular ways, whatever the reasons, such data can be generally used. Thus, this is sufficiently relaxed, but the data for several countries, not too many, are not suitable for the usage of our “forecasting tool”.

### Forecasting the spread

With these reservations, the first point of maximum *t*_*top*_ of *u*(*t*) is a good estimate for the duration of the epidemic when the “hard” measures are used and the response is “hard”. If the response to total number of infections is “soft”, the *w*–function is supposed to be used. This is assuming that the measures are systematic and the focus is on the detection and isolation of infected people.

We note that the approximate reflection symmetry of *du*(*t*)/*dt* for the *u*(*t*) in the range from *t* = 0 to *t*_*top*_ can be interpreted as *Farr’s law of epidemics under aggressive management*. Generally, the portions of the corresponding graph before and after the *turning point* are supposed to be essentially symmetric to each other. This is not exactly true for *du*(*t*)/*dt*, the first half is a bit shorter than the second. See Figure 2 and the others; the turning point is at max {*du*(*t*)/*dt*}. For *w*(*t*), the period after the turning point is somewhat longer than that before.

**Figure 1.**
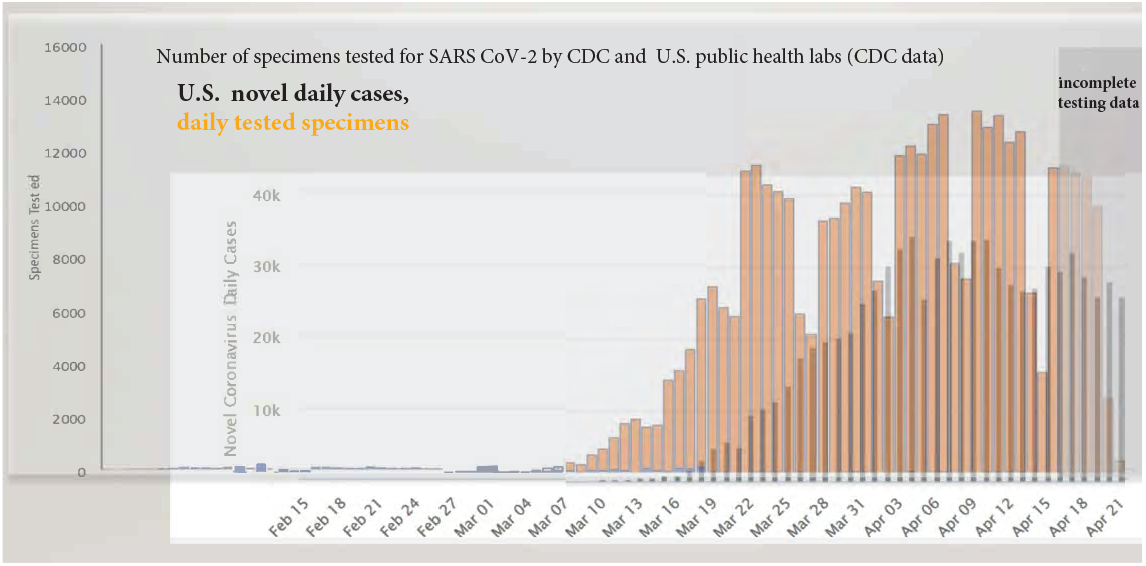
Daily testing vs. new daily cases for USA

**Figure 2.**
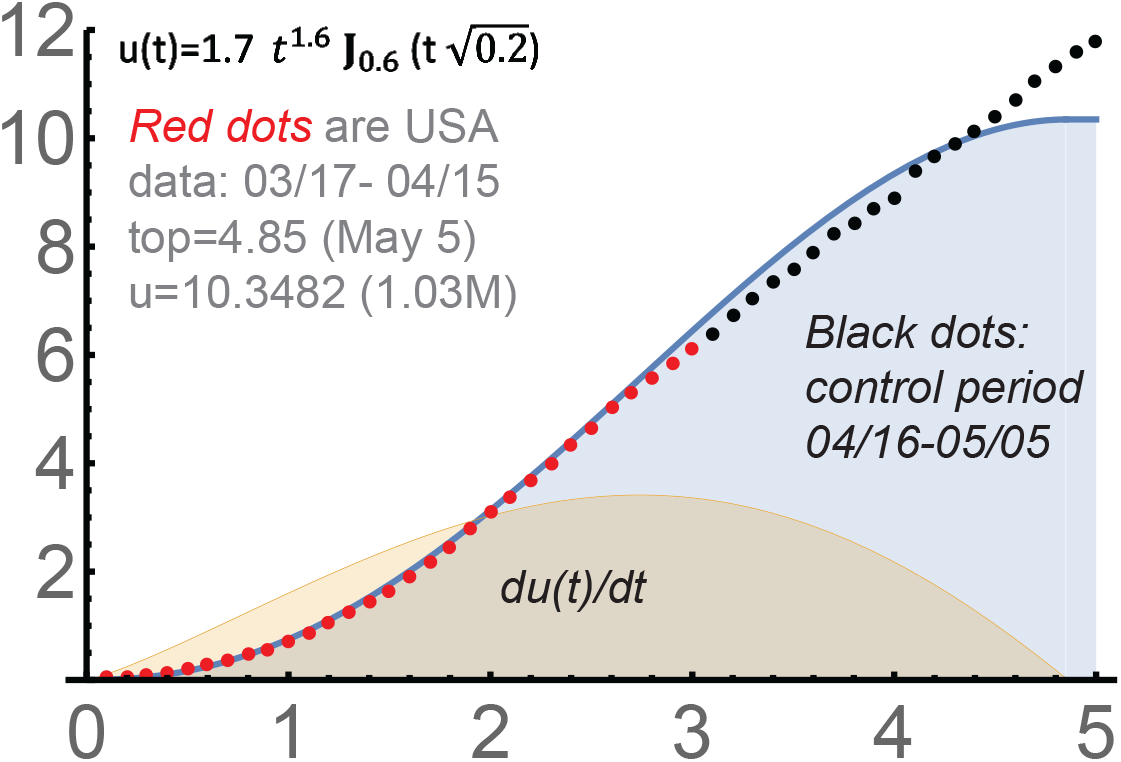
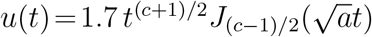 for *c* = 2.2, *a* = 0.2

**Figure 3.**
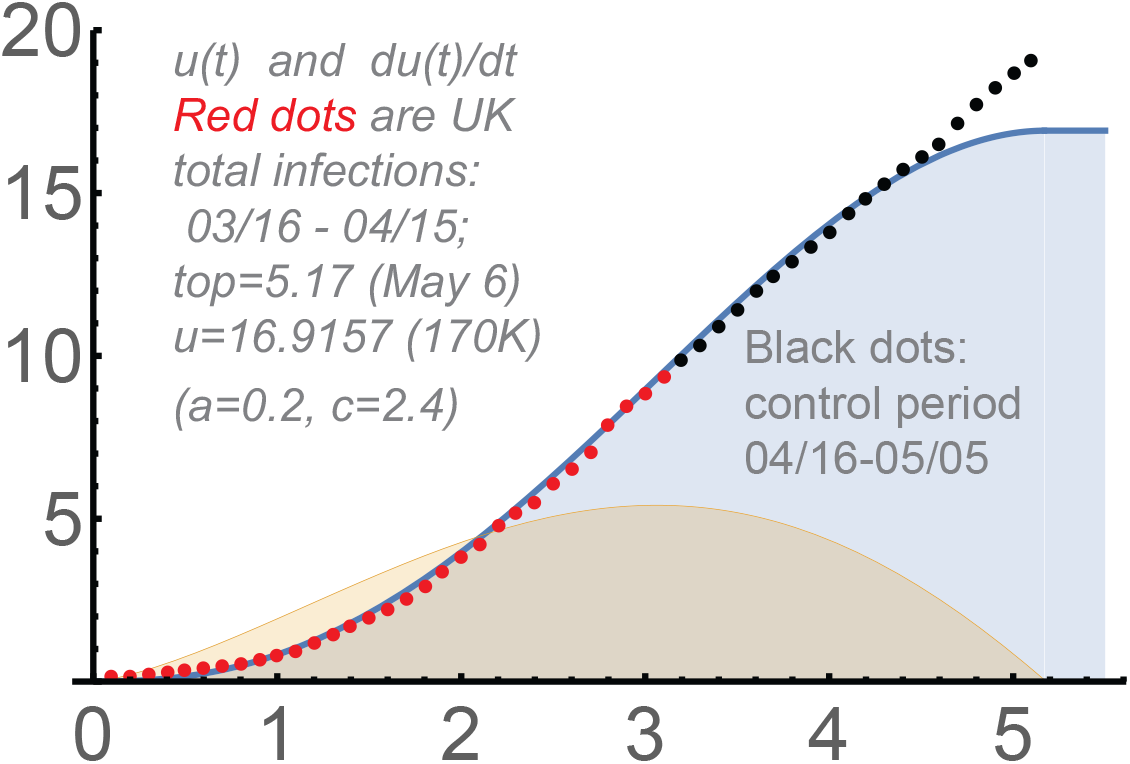
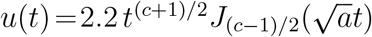 for *c* = 2.4, *a* = 0.2

After the saturation is reached, the second phase begins. The formula is *Ct*^*c*/2^ cos (*d* log (*t*)), where *t* is time, *d* reflects the intensity of “soft” measures, and *c* is the initial transmission rate. Here *c*/2 ≈ 1, so the number of new daily detected cases is essentially constant. If *C* is small enough, the spread can be controlled: new clusters of the disease can be then promptly detected, an so on. Also, asymptomatic (mild) cases begin to dominate at the second phase, which is a positive development too. Mathematically, the first phase based on “hard” measures is necessary for phase 2.

As any model, our one is based on various simplifications. We assume that the number of people perceptive to the virus is unlimited, i.e. we do not consider epidemics with the number infections comparable with the whole population. Also, we do not take into consideration the average durations of the disease the quarantine periods in our model. The *total* number of *detected* infections is what we are going to model, which is commonly used and systematically reported. In spite of all these simplifications, our 2-phase formula works very well.

We mention here a strong connections with *behavioral finance* : *momentum risk-taking* from [7]. Practically the same *u*(*t*) as above serves “profit taking” in stock markets. The initial polynomial growth of *u*(*t*) is parallel to the “power law” for share prices; *w*(*t*) and our solution for the *phase 2* occurred there as well

## 2. Two kinds of management

There is a long history and many aspects of mathematical modeling epidemic spread; see e.g. [1] for a review. We restrict ourselves only with the dynamic of *momentum managing epidemics*, naturally mostly focusing on the middle stages, when our actions must be as precise as possible. The two basic modes we consider are essentially as follows:

A. aggressive enforcement of the measures of immediate impact, where testing-detection-isolation is the key, reacting to the current *absolute* numbers of detected infections, i.e. in the hardest possible way;
B. a more balanced and more defensive approach when mathematically we react to the *average* numbers of infections to date and the employed measures are of more indirect and palliative nature.

### Hard and soft measures

The main examples of (*A*)–type measures are prompt detection and isolation of infected people and those of high risk to be infected, and closing places where the spread is the most likely. Actually the primary measure here is *testing* ; the number of tests is what we can really implement and control. The *detection* of infected people is its main purpose, but the number of tests is obviously not directly related to the number of detections, i.e. to the number of *positive tests*. The efficiency of testing requires solid priorities, focus on the groups with main risks, and solving quite a few problems.

Even simple mentioning problems with testing, detection and isolation is well beyond our article. However, numerically we can use the following. During the epidemics, essentially during the stages of linear growth, which are the key for us, the number of positive tests can be mostly assumed a stable fraction of the total number of tests. This is demonstrated in Figure 1. Such proportionality can be seen approximately from March 16 in this figure.

The measures like wearing protective masks, social distancing, recommended self-isolation, restricting the size of events are typical for (*B*). They are aimed at reducing *c*, which heavily impacted the differential equations we obtained in [6]. However the main difference between the modes, (*A*) vs. (*B*), is the way the number of infections is treated: the *absolute* number of infections is the trigger for (*A*), whereas the *average* number of infections to date is what we monitor under (*B*). The latter results in a relatively slow response; we wait until the *averages* reach some levels.

The mode (*AB*) from [6], a combination of “hard” measures with “soft response”, is important at later stages of *Covid-19*.

### Hard measures are the key

Generally, the (*A*)–type approach provides the fastest and hardest response to the changes with the number of infections. We somewhat postpone with our actions until the averages reach proper levels under (*B*), and the measures we implement are “softer”. Mathematically, the (*B*)-mode is better protected against stochastic fluctuations, but it is slower and generally cannot alone lead to the termination of the epidemic, which we justify within our approach.

The main objective of any managing epidemics is to end them quickly. However the excessive usage of hard measures can lead to the recurrence of the epidemic, some kind of “cost” of our aggressive interference in a natural process. Unless *herd immunity* is a possibility, this can be avoided only if we stick to the prevention measures as much as possible even when the number of new infections goes down significantly. Reducing them too much on the first signs of improvement is a way to the recurrence of the epidemic, which we can see mathematically.

### Some biological aspects

The *viral fitness* is an obvious component of the transmission rate *c*. Its diminishing over time can be expected, but this is involved. This can happen because of the virus replication errors. The *RNA* viruses, *Covid-19* included, replicate with fidelity that is close to error catastrophe. See e.g. [8] for some review and predictions. Such matters are well beyond this paper, but one biological aspect must be mentioned, concerning the *asymptomatic cases*.

The viruses mutate at very high rates. They can “soften” over time to better coexist with the hosts, though fast and efficient spread is of course the “prime objective” of any virus. Such softening can result in an increase of asymptomatic cases, difficult to detect. So this can contribute to diminishing *c* we observe, though not because of the actual decrease of the spread of the disease. We model the available (posted) numbers of total infections, which mostly reflect the symptomatic cases.

To summarize, it is not impossible that the replication errors and “softening the virus” may result in diminishing *c* at later stages of the epidemic, but we think that the reduction of the contacts of infected people with the others dominates here, which is directly linked to behavioral science, sociology and psychology.

## 3. The Bessel-type formulas

We will need the definition of the *Bessel functions* of the first kind:

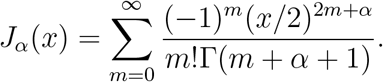

See [9] (Ch.3, S 3.1). Practically, use `BesselJ` [*α*, `x`] in Mathematica.

The key point is that measures of type (*A*) have “ramified” consequences, in some contrast to (*B*). Namely, isolated infected individuals will *not* transmit the virus to many people, they will not infect many others and so on; thus the number of those protected due to a single isolation grows over time. Combining this with our approach to the power law of epidemics, we arrive at the differential equation for the total number of detected infections *u*(*t*), *w*(*t*):

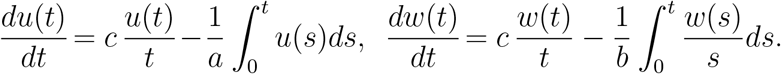

The fundamental solutions are 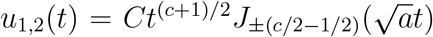 and 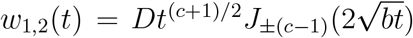 for some constants *C, D*. Note that both are ∼ *t*^*c*^ for small *t*. Unless stated otherwise we use *u* = *u*_1_ and *w* = *w*_1_, *the dominant solutions*. Actually, *u*_1_ and *u*_2_, both, are necessary, as we will see.

There is a surprisingly perfect match of the *total* number of infections for *Covid-19* during the initial and middle stages with our *u*(*t*), from the moment when these numbers begin to grow “significantly” and during some time after the “turning point”. In the USA and UK, it certainly was from March 15 till April 15. Epidemics are very stochastic processes, so such a precision is unexpected.

Let us mention that https://ourworldindata.org/coronavirus is mostly used for the data, updated at 11:30 London time. *We always set x* = *days/*10 *in this article*.

### USA in phase one

The scaling coefficient 1.7 in Figure 2 is adjusted to match the real numbers. For the USA, we set *y* = infections/100*K*, and take March 17 the beginning of the period of “significant growth”. The parameters are *c* = 2.2, *a* = 0.2.

The *dots* show the corresponding actual *total* numbers of infections. The *red dots* were used to obtain the formula; they *perfectly* match 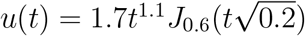 in Figure 2. The initial projection was: *u*_*top*_ = 10.3482 at *t*_*top*_ = 4.85 (48.5 days from 03/17), i.e. with about 1*M* infections at May 5. The total number of infections was 609516 at 04/15. The projection was of course a under the assumption that the USA would follow the way of Europe, except for Belarus, Russia, Sweden, and UK (at that time).

The *black dots* show the test period, which was till *t*_*top*_, May 5. Mathematically, the assumption was that *the intensity of hard measures would continue to be proportional to the total number of detected infections to date*. However it was the case only till the end of April. If *u*(*t*) is not applicable, as for the USA and UK, then the usage of *w*(*t*) is supposed to be the next natural step according to [6].

Even if *t* = *t*_*top*_ is reached, this is not the end of the epidemic. The data from South Korea and other countries that went through the “saturation”, demonstrate a period of essentially linear growth of the total number of cases around and after *t*_*top*_, which is our *phase 2*. More exactly, *u*(*t*) ∼ *t*^*c*/2^ cos (*d* log (*t*)) during this phase.

### Covid-19 in UK

The *red dots*, when the formula was obtained, are from 03/16 till 04/15; 18 must be added to the *y*-values of our “red dots”, the initial number of cases at 03/16, to match the actual total numbers. The *black dots* constitute the control period: 04/16-05/05.

Now *c* = 2.4, *a* = 0.2 work fine, and the scaling coefficient is 2.2. The total number of cases is divided by 10*K*, not by 100*K* as for the USA. The “*u*–saturation moment” was 5.17, i.e about 51 days after March 16, somewhere around May 6. The estimate for the corresponding number of infections was about 170*K*, with all ifs. This was assuming that the “hard” measures would be employed at the same pace as before April 15, i.e. following mode (*A*), the most aggressive approach.

### Sweden 03/07-04/23

This is an example of the country that remains essentially “open”. Actually, they actively do testing of infected people, a “hard” measure from our perspective. Also, the strength of the health-care in this country must be taken into consideration, and that Sweden is surrounded by the countries that fight *Covid-17* aggressively. The growth of the total number of cases was essentially quadratic for a relatively long period, which is what “power law” states for the epidemics with minimal “intervention”. By now, the growth is linear; the reduction from *t*^*c*^ to *t*^*c*/2^ follows from our theory. Mathematically, our *u*– formula is still applicable, but for low *a* = 0.1. Here *y* is the total number of cases divided by 1000 ; add 137, the initial value, to our *y*. The projected *u*–saturation was around May 17, with all standard reservations; see Figure 4. Of course the usage of *u, w* is questionable for the countries that do not employ hard measures.

**Figure 4.**
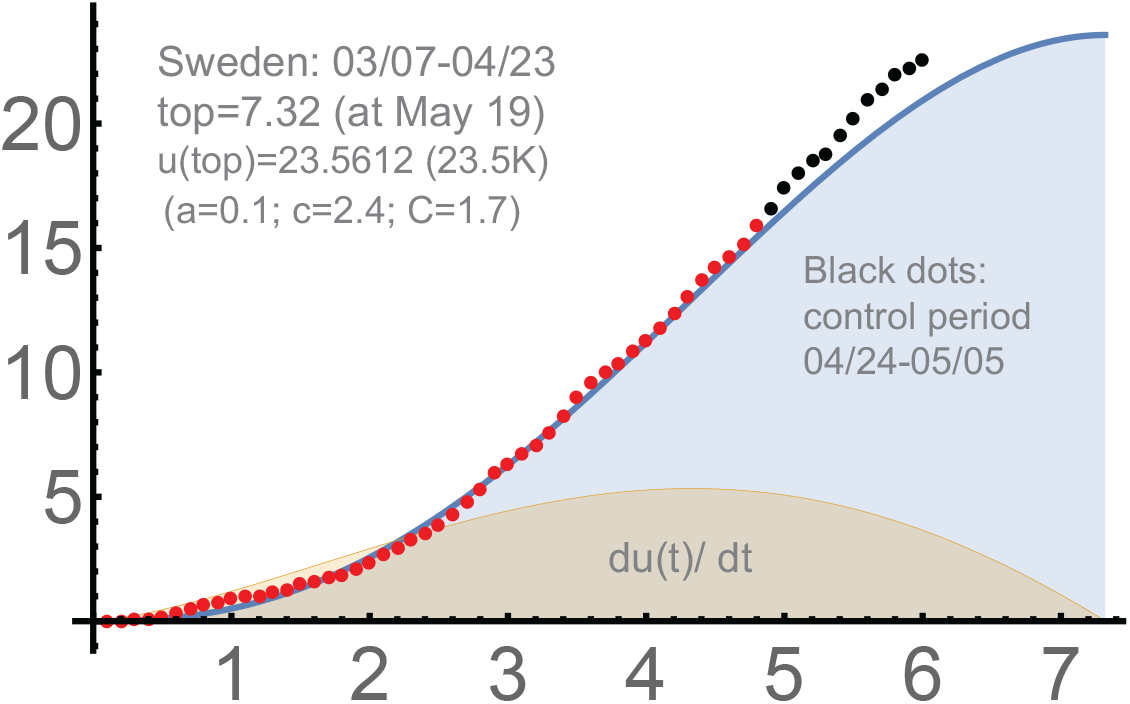
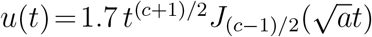 for *c* = 2.4, *a* = 0.1

### Israel: “saturation”

The country went through the “saturation”. Israeli population is diverse, which has a potential of significant fluctuations of the number of cases and various clusters of infection. However its solid response to *Covid-19* and good overall health-system, made the growth of the spread sufficiently predictable. We divide the total number of cases by 1000, as for Sweden; see Figure 5.

**Figure 5.**
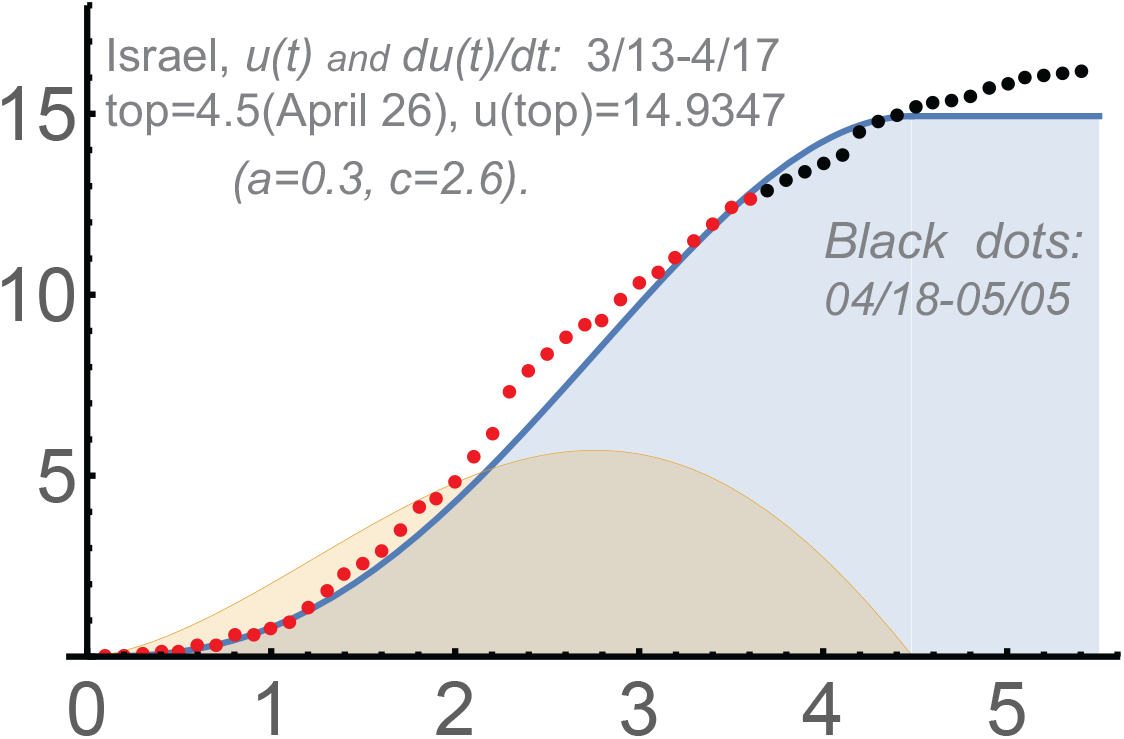
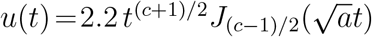 for *c* = 2.6, *a* = 0.3

**Figure 6.**
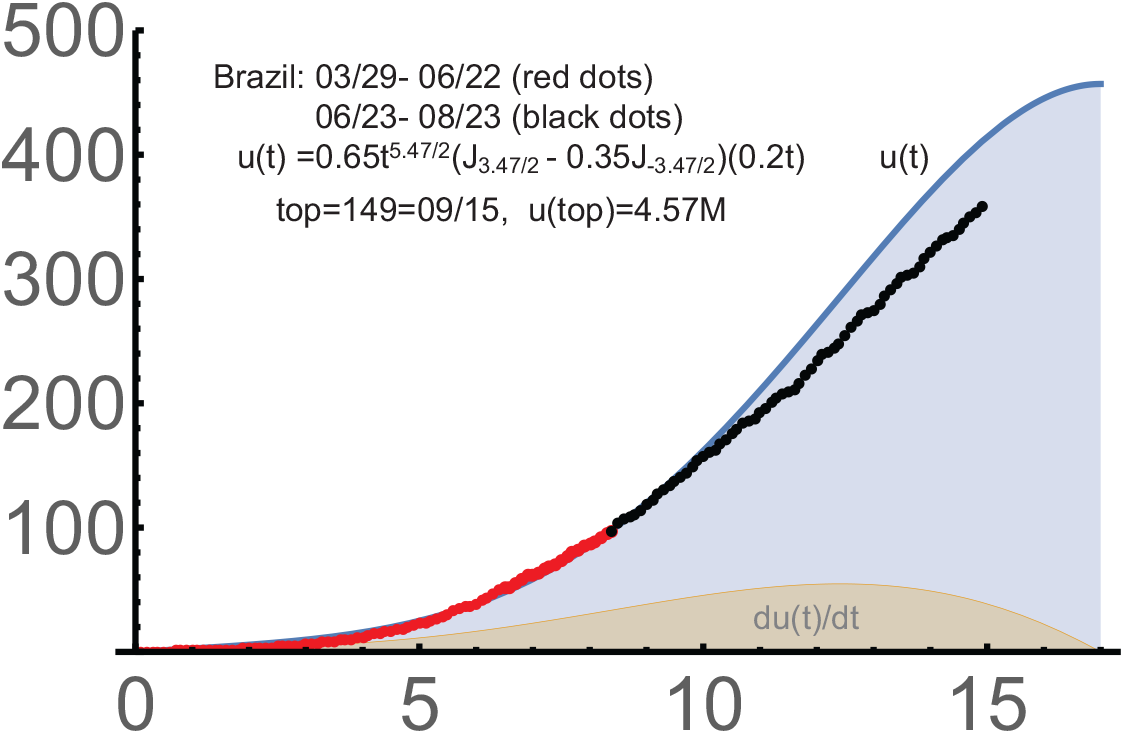
Brazil: *c* = 4.47, *a* = 0.04, *C* = 0.65.

The *red dots* began March 13, when the *total* number of detected infections was 96, and stopped April 17; the remaining period till May 5, shown by the black dots, was the “control one”. The saturation forecast went through almost perfectly, but there were significant fluctuations in process. After April 26, the predicted moment of the saturation, the growth of the total number of (known) infections is supposed to be mild linear and became so. The parameters are: *a* = 0.3, i.e. the intensity of hard measures is better than with the USA, UK, and *c* = 2.6. The latter means that the initial transmission coefficient was somewhat worse than those in the USA and UK, possibly due to the greater number of “normal” contacts. Recall that *a, c* are parameters of our theory, related to but not immediately connected with the real factors.

The “bulge” in the sequence of *red dots* can be essentially “fixed” using the second Bessel-type; this is quite similar for Brazil, Italy, Japan, Germany considered below.

### Brazil and India

There are ongoing processes in both countries, which are actually similar mathematically. Phase 1 can be very well modeled using the combination of two our solutions *u*_1_,*u*_2_. We take them here and below with the same scaling coefficient *C*.

They parameters are *a* = 0.04, *c* = 4.47, *C* = 0.65 for Brazil, where we set *y* =cases/10*K*. In contrast to the previous examples, we use *u*_1_ = *Cu*^1^ *and* the second (non-dominant) solution *u*_2_ = *Cu*^2^:

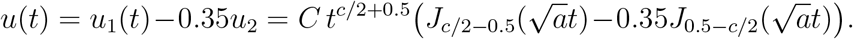

Note that the parameter *a*, the intensity of “hard measures”, is very low, and *c* is quite large.

The parameters are even more extreme for India: *a* = 0.035, *c* =

5.2. This country is still in the period of polynomial growth of the spread, as of August 25. The analysis of India is obviously important to understand the future of *Covid-19*. We provide the corresponding *u*– curve. Recall that it is under the assumption that the “hard” measures are used in a steady manner, which was the case in almost all Western Europe, China, South Korea, the USA (during some periods), and in several other countries. Also, a sufficiently high level of healthcare system and the uniformity of measures in the whole country are necessary for the success of such measures.

Even when all these factors are present, the spread can be very intensive. This occurred in the USA, where the *u*-curve and *w*-curve served well only the middle stages. In India, the parameters of the spread were the most extreme among the countries we consider in this paper:

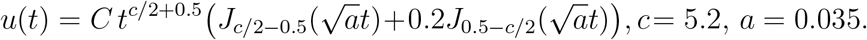

We set *y* =cases/10*K* in Fig. 7. The period we used to determine the parameters was 3/20-8/23 (no “black dots”). The starting number of detected cases was 191, which was subtracted,

**Figure 7.**
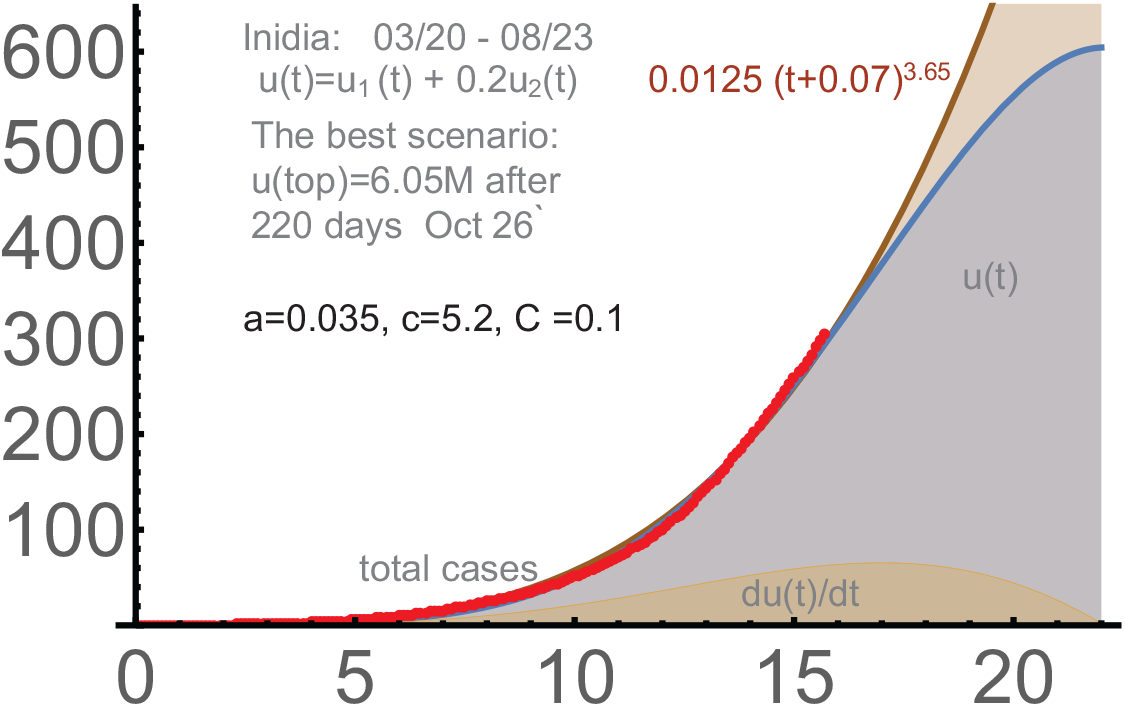
*India* : 3/20 − 8/23, *c* = 5.2, *a* = 0.035, *C* = 0.1.

Actually, 0.0125 (*t*+0.07)^3.65^ gives a good approximation for the main part of the period; see Figure 7, where the graph of this function is shown as brown. So this is really a period of (strong) polynomial growth, but *obviously* not of any exponential growth. We never observed exponential growth of the number of detected infections with *Covid-19* beyond some very short initial periods.

Let us update Figure 7 till 10/07/2020. The parameters there were determined *before* the turning point, so they were not that reliable. As of the beginning of November, India presumably reached this point. Interestingly, only *c* required some adjustment; it is now *c* = 5.75 vs *c* = 5.2 determined for the period till 8/23. *All other coefficients, including the exponent are unchanged*. The same (brown) power function 0.0125 (*t* + 0.07)^3.65^ provided a good approximation till the turning point. The match with our Bessel-type solution was almost perfect, though of course much will depend on the further developments in India. Recall that *y* =cases/10*K* in Fig. 8. The red dots are those used in Figure 7 (till 8/23). The clear polynomiality of the curve of total cases in India and the match with our *u*-functions are of course important confirmations of our theory. We have:

**Figure 8.**
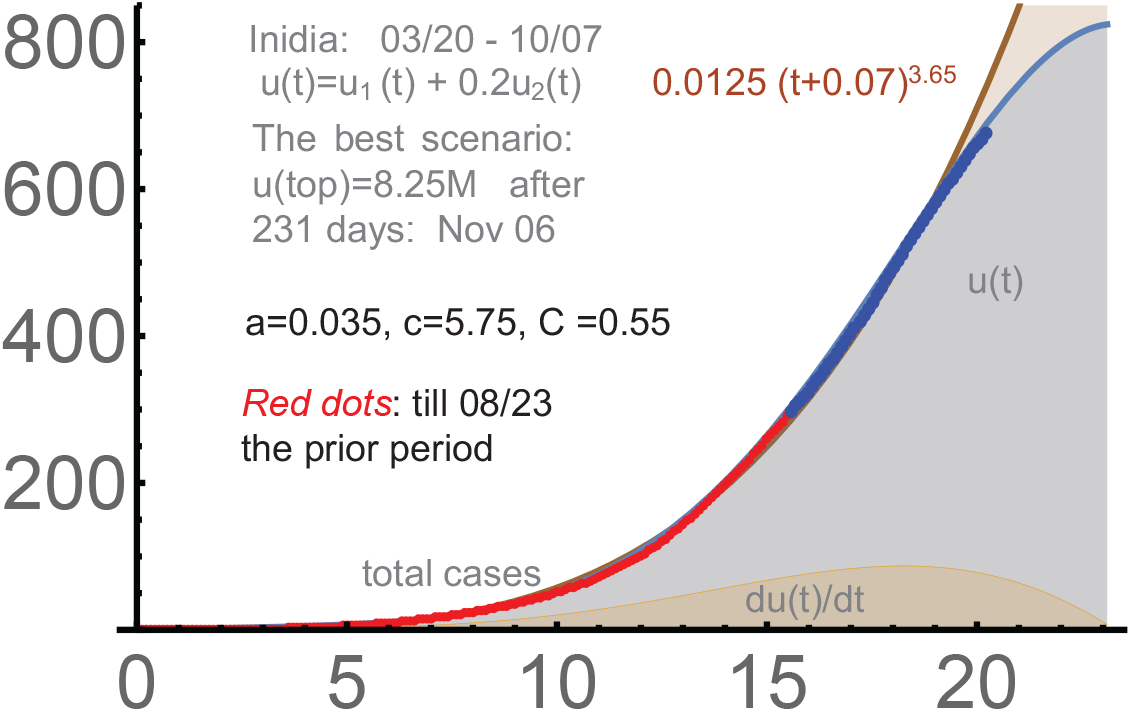
*India* : 3/20 − 10/7, *c* = 5.75, *a* = 0.035, *C* = 0.55.

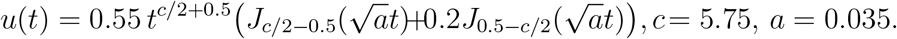

## 4. Forecasting cones

Our function *u*(*t*) models well the period of intensive growth when the hard measures are coupled with the most aggressive respond to the current number of total cases, i.e. under (*A*)-mode. Then *t*_*top*_, the first zero of *du*(*t*)/*dt* is a reasonable estimate for the “phase-one saturation” in almost all Europe, about 20 states in the USA and other places.

Reducing hard measures (if any) almost after the turning point, or after what looks like a turning point naturally influences such forecasts. The growth can be expected linear then, but the number of daily new infections can be very high. From our perspective, this means a switch from mode (*A*) to mode (*B*) or (*AB*). With the latter, the hard measures are still present, but the response becomes softer, of type *B*. For instance, if the number of new cases is essentially a constant, even uncomfortably high, the (*B*)–response is to keep the testing-detection constant too. Type– (*A*) response will be “parabolic” in this case.

Obviously the improvements with testing and better capacities for isolation and treatment is quite a consideration for reopening places with potentially high risks of the spread of *Covid-19*. Also, people who suspect that they are infected request help begin more actively at this stage, which works in the same direction as any “hard” measures.

Mathematically, if the new daily cases are constant but relatively high, a different kind of modeling is required, which is (*AB*). Our theory provides this. The assumption is that the hard measures are still in place, but the response to the current total number of detected infections is via *U*(*t*)/*t* instead of *U*(*t*); see Section 2.

This model is governed by *w*(*t*). The point 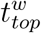 *>*where the (first) max*>*imum of *w*(*t*) occurs seems a reasonable upper bound for the “technical *>*saturation”. The prior 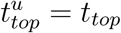 then is a lower bound; we obtain some *>forecast cone* : between 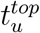 and the *w*-curve.

Actually even relatively minor deviations with *a, c* can lead to significant changes of *u*(*t*) over time, so there are some natural “cones”. However, we have something more fundamental here. The switch to *w*(*t*) is due to a different kind of management.

When the daily numbers of new cases are high, there are of course significant chances of new clusters and fluctuations of the data of all kinds. So we need to model a process with many uncertainties. However, we think that this is basically no different from what we did for the middle stages, where *u*(*t*) was surprisingly efficient.

Recall that 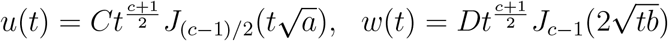

### The cone for the USA

The prior 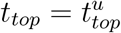 *>*for *u*(*t*) was May 5, with *a, c, C* calculated on the bases of the data till April 16. It was under the expectations that the “hard” measures would applied as in (*A*) (as before April 16). However the presence of 3 major spikes in the daily cases and other factors like *de facto* relaxing hard measures obviously significantly delayed the saturation.

Whatever the reasons, the switch to the (*AB*)-mode and *w*(*t*) was for us a natural adjustment. However, it appeared insufficient for the USA. We provide the *blue dots*, obtained after the forecast cone was determined. For Sweden, the cone was not expected to work, but the dots for the USA appeared following a kind of “Sweden pattern”.

The cone is defined as the area between *u*(*t*) extended by a constant *u*(*t*_*top*_) for 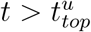 and the graph of *w*(*u*) till 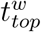, which is approximately May 30, 2020. The parameters *b, D* of *w*(*t*) are calculated to ensure good match with *red dots*; *c*, the initial transmission rate, is the same for *u*(*t*) and *w*(*t*). The graphs of *u*(*t*) and *w*(*t*) are very close to each other in the range of *red dots*. The above relation for *C/D* holds with the accuracy about 20%.

We mostly monitor the *trend*, the “derivative” of the graph of black dots, which is supposed to be close to the derivative of *w*(*t*) or *u*(*t*). See Figure 9. The spikes with number of cases are acceptable if the dots continue to be “parallel” to *u*(*t*) or *w*(*t*) (or in between).

**Figure 9.**
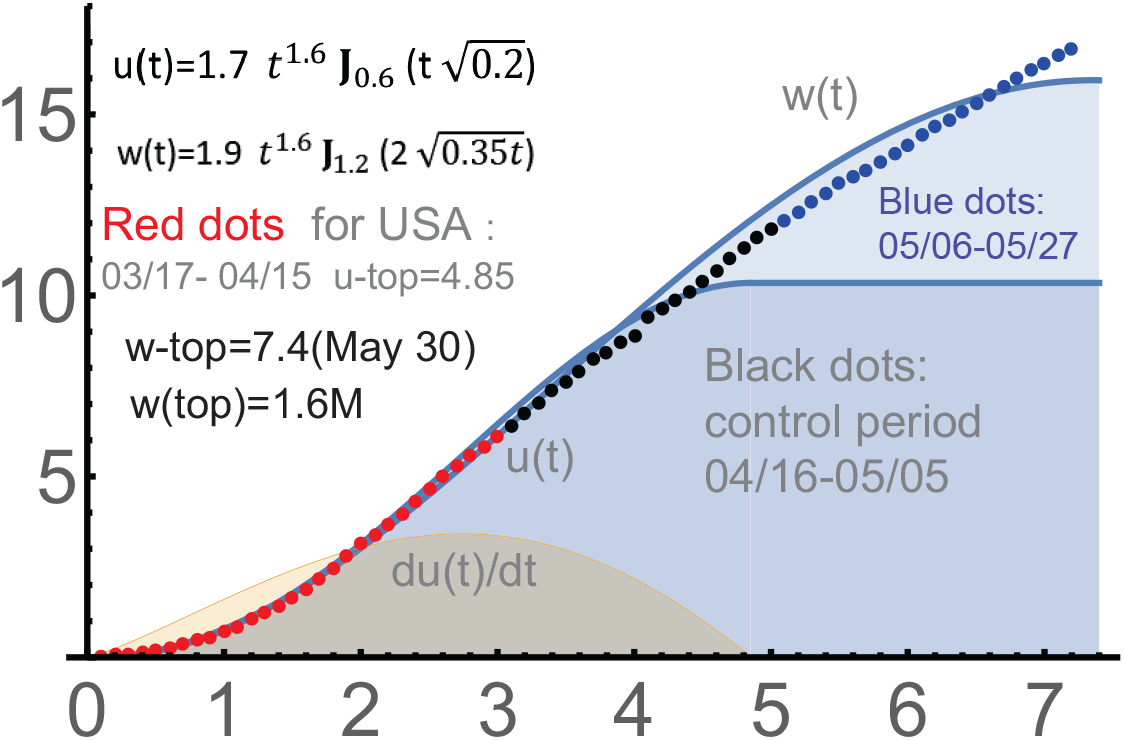
USA: *c* = 2.2, *a* = 0.2, *C* = 1.7; *b* = 0.35, *D* = 1.9.

### UK and Sweden

The graphs of *u*(*t*), *w*(*t*) with red-black-blue dots obtained then and now for UK are in Figure 10. The expected 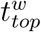 was around 06/10 with the total number of detected infections about 330K. These “predictions” went through; UK is now in phase 2: Figure 17.

**Figure 10.**
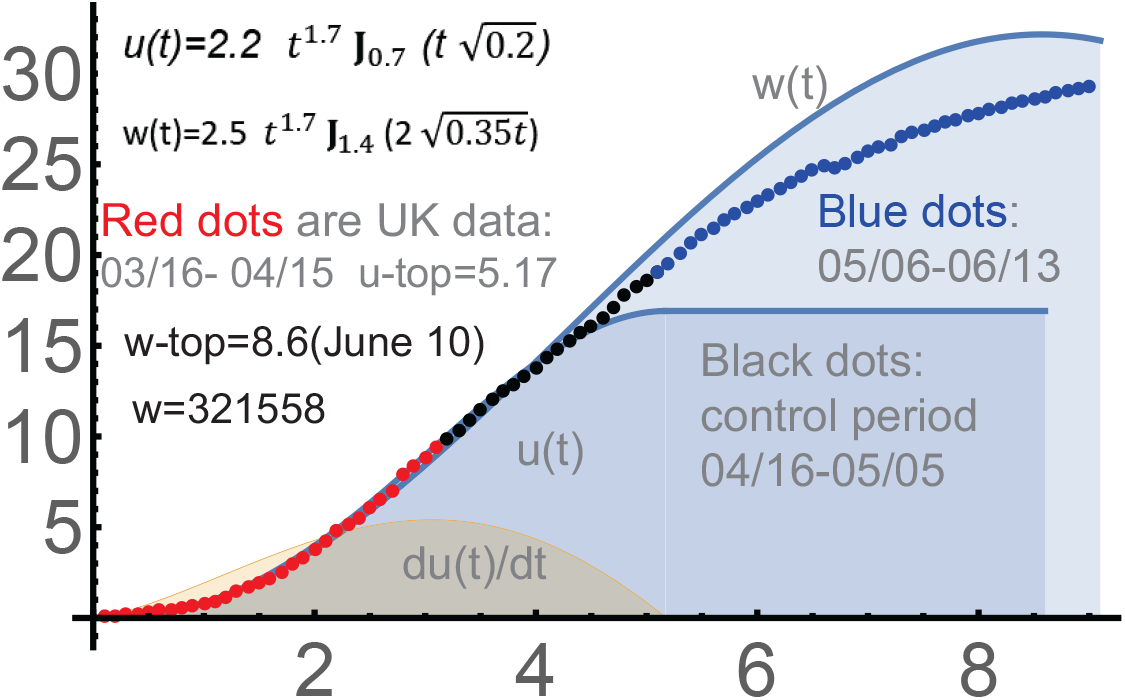
USA: *c* = 2.4, *a* = 0.2, *C* = 2.2; *b* = 0.35, *D* = 2.5.

**Figure 11.**
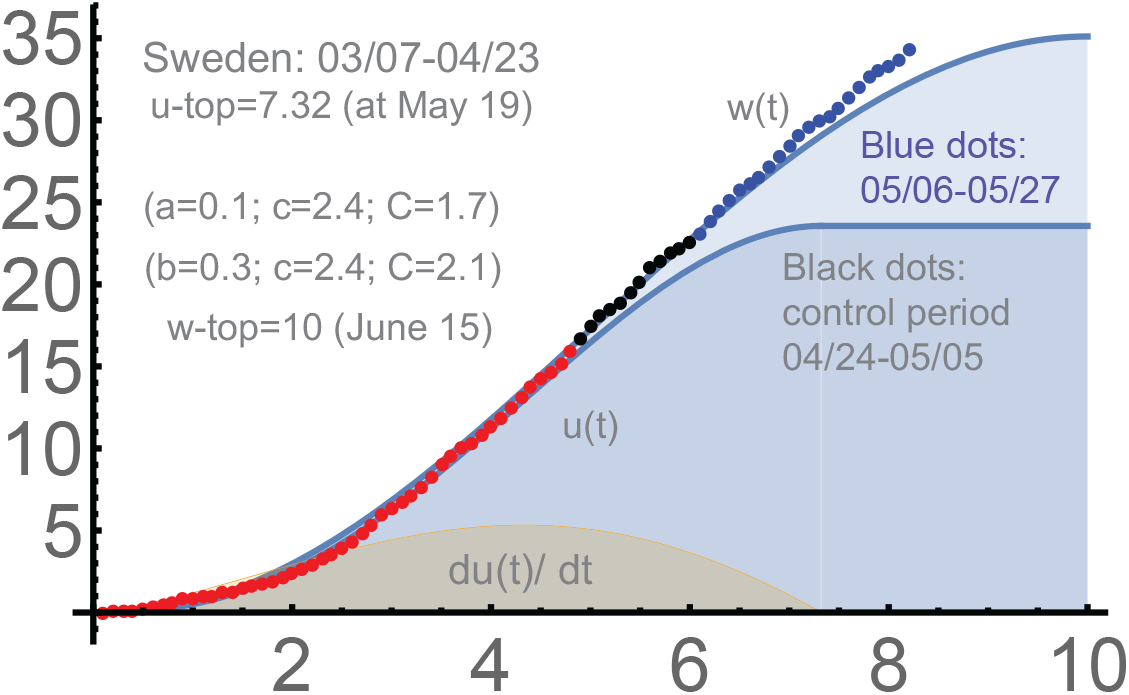
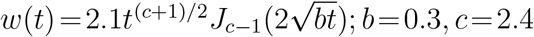.

**Figure 12.**
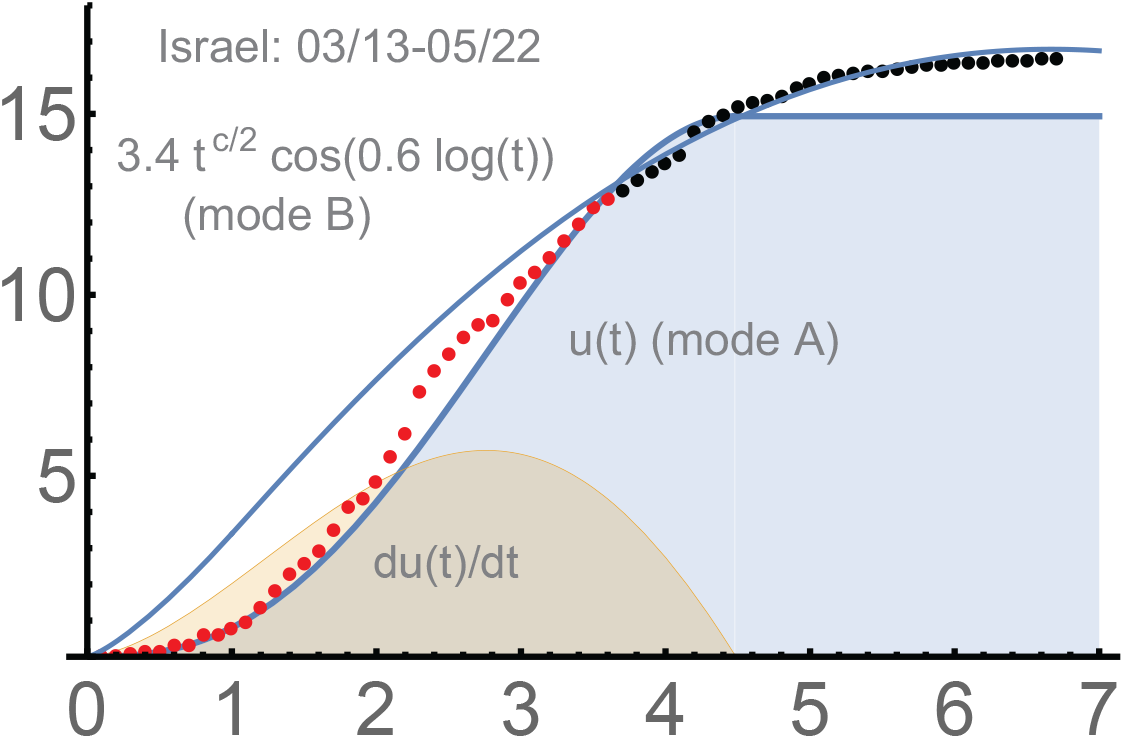
Israel: *c*=2.6, *a*=0.3, *d*=0.6

**Figure 13.**
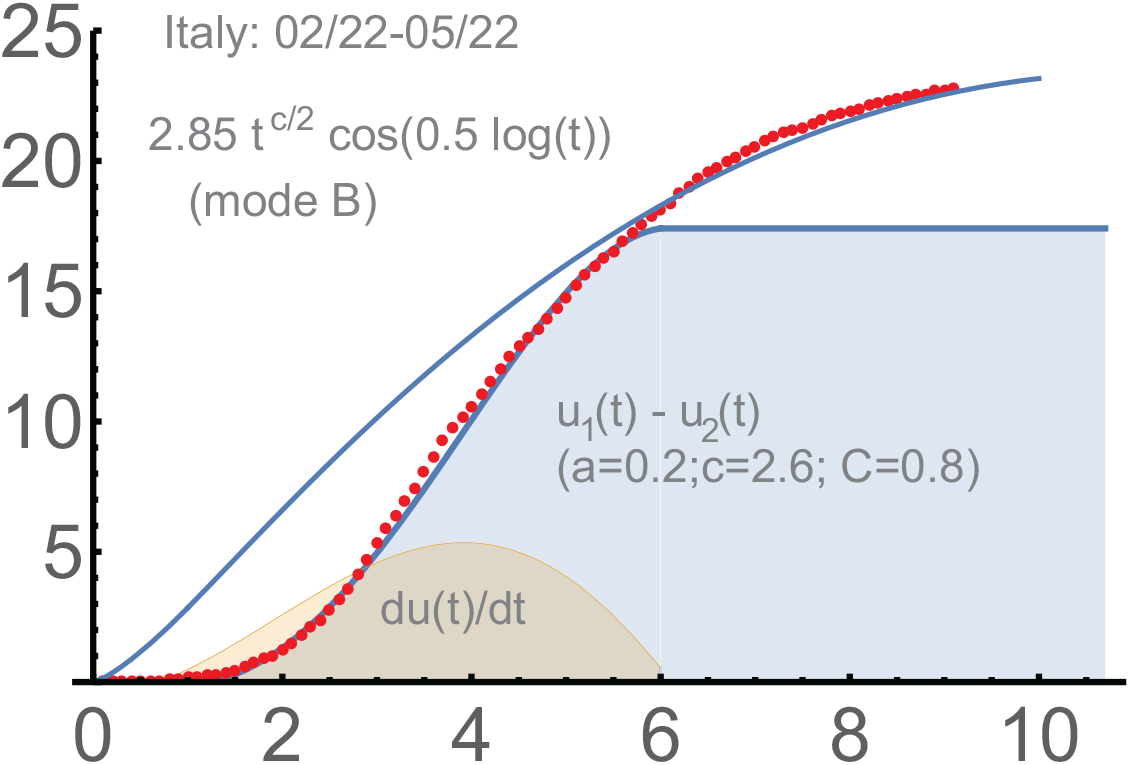
Italy: *c* = 2.6, *a* = 0.2, *d* = 0.5

**Figure 14.**
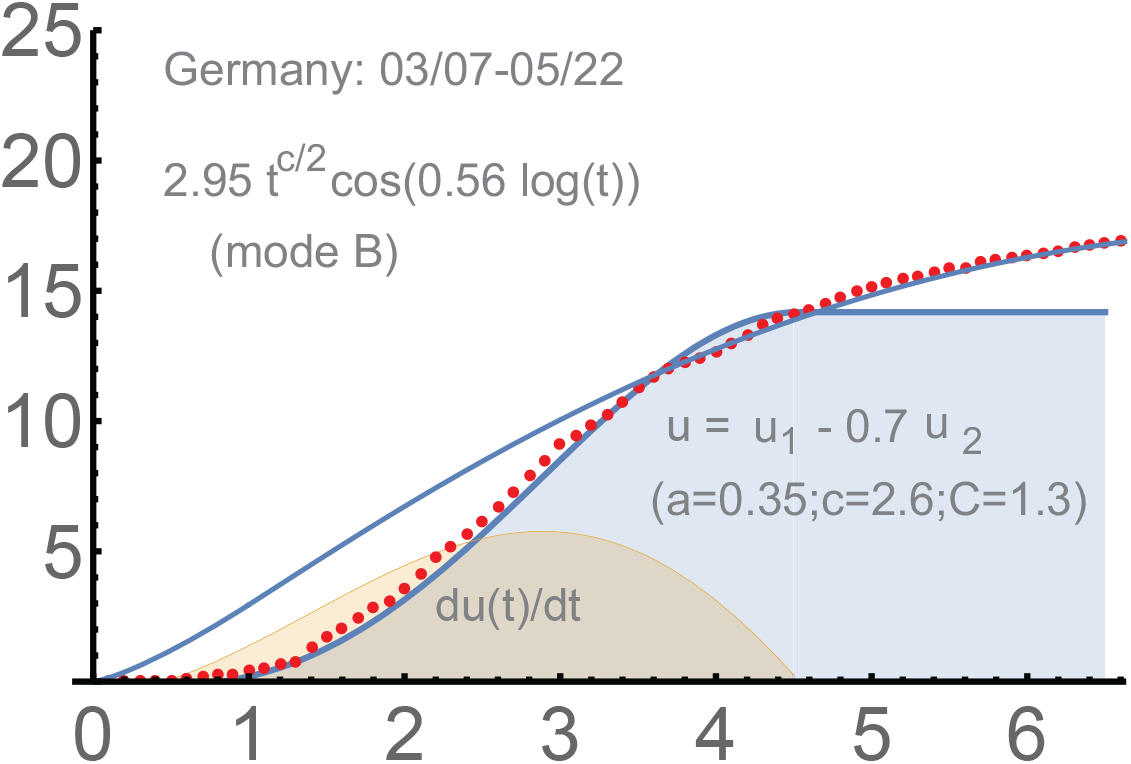
Germany: *c* = 2.6, *a* = 0.35, *d* = 0.56

**Figure 15.**
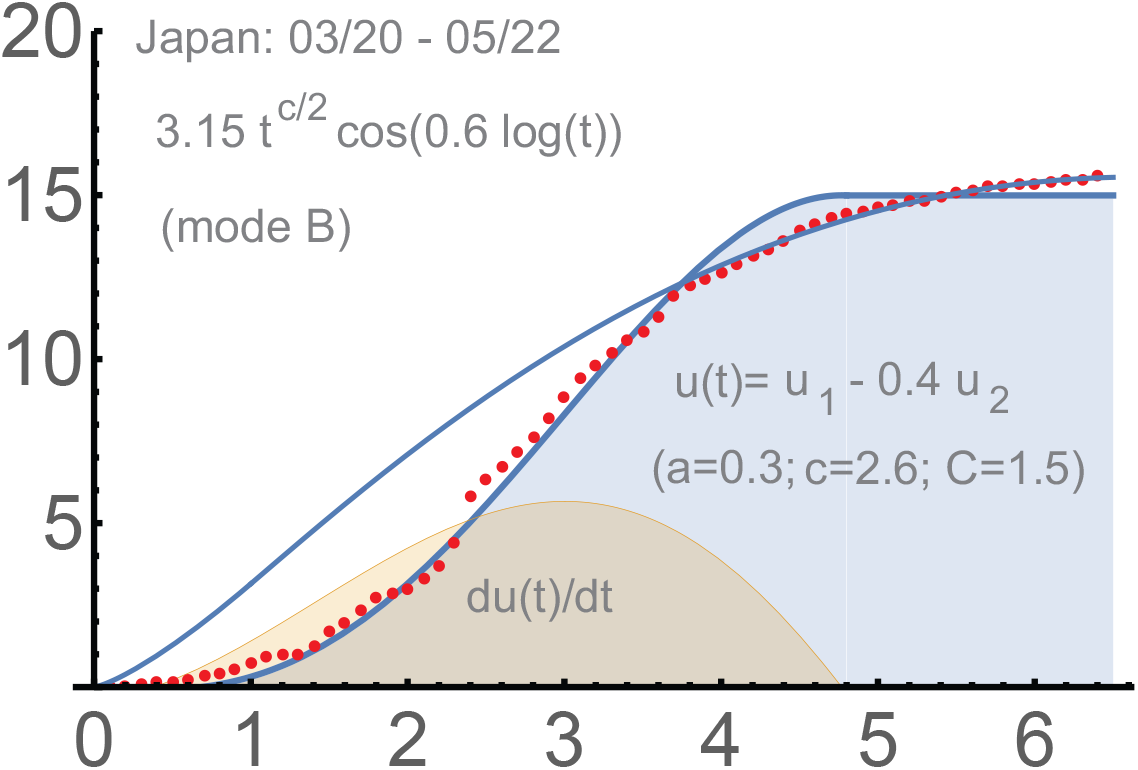
Japan: 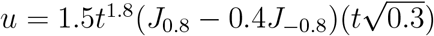.

**Figure 16.**
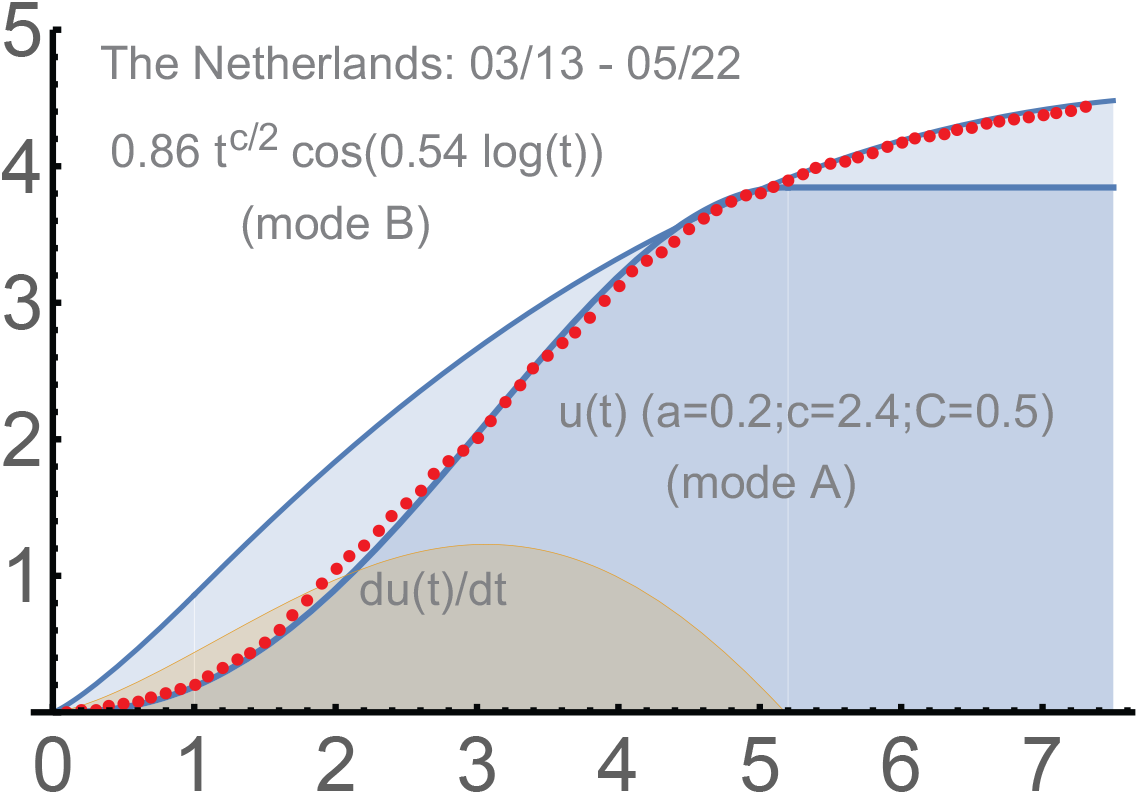
The Netherlands: 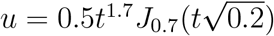.

**Figure 17.**
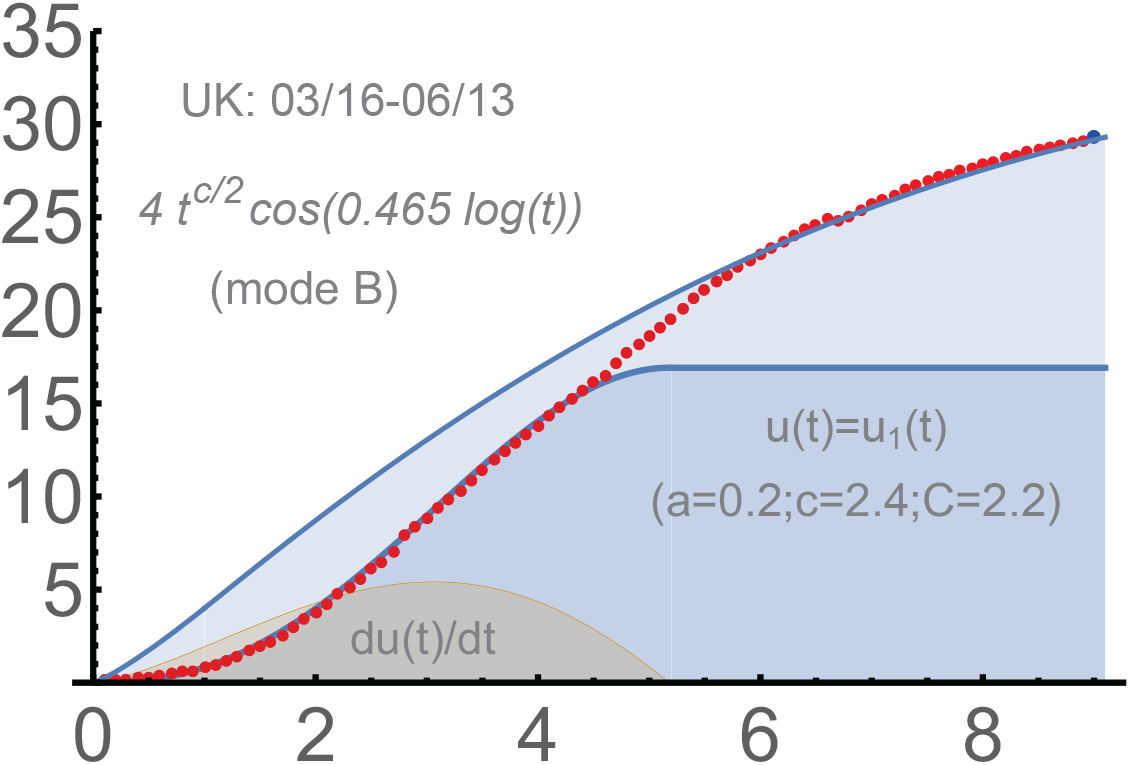
UK: 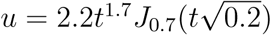.

For Sweden, the total number of infections in Sweden was actually *not supposed* to match our *u, w*–curves, because this country does not follow hard ways of fighting *Covid-19*. This can be clearly seen in Figure 11. We note that the USA and UK (for some time) have followed the “Sweden approach”, relaxation hard measures *on the first signs of improvements*, when the numbers of new daily cases were essentially constant but still very high.

## 5. Two-phase solution

Let us now focus on our *two-phase* model for the number of total infections, which is from the beginning of the intensive spread through the saturation *t*_*top*_ of type (*A*), and then (phase 2) till the “final saturation” under the formulas for the (*B*)–phase. It works surprisingly well for the countries that reached phase 2, so the (*AB*)–mode was mostly unnecessary for them.

The corresponding solution for phase 2 is in Section 6 from [6]. It is

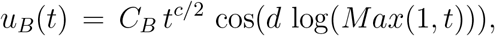

where *c* must be the one we used in.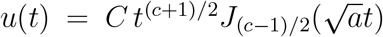 We determine here the parameters *a, c, d*, and the scaling coefficients *C, C*_*B*_, using the whole period till May 22 (2020), unless for Israel, where *a, c, C* were found in the middle of April, and for UK.

Here *Max*(1, *t*) is to avoid some ambiguity at *t* = 0; and we use *u*_*B*_ anyway for *t* > 1. Importantly, we start *u*_*B*_ at *t* = 0, not at some intermediate point. Practically, this means that the (*B*) formula we find provides the rest of the curve after *t*_*top*_. After that moment, the usage of “soft” measures can be mathematically sufficient to end the epidemic. Note that *d* can be determined only in the vicinity of *t*_*top*_, when/if it is reached.

The epidemic is of course far from over, but the first “cycle” hopefully approaches the end in these countries; so no test periods, “blue dots”, seem necessary. Recall that the parameters *a, c, C* for the first phase are mostly obtained on the basis of the period before or around the “turning points”; the same *c*, the initial transmission coefficient, is supposed to be used for both, (*A*) and (*B*), according to our theory.

For Israel, the period was 03/13-05/22. Here *c* = 2.6, *a* = 0.3, *d* = 0.6. The scaling coefficients are *C* = 2.2, *C*_*B*_ = 3.4. Note that 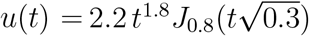 is the one we used in Figure 5.

### Italy: 2/22-5/22

See Figure 13. The starting point is 2/12, when the total number of infections was 17; in this paper, we always subtract this initial value when calculating our *dots*. One has:

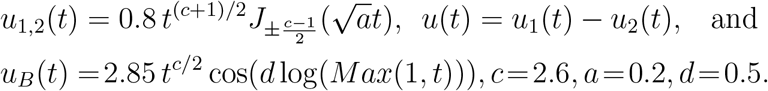

Here we use the second, non-dominant, solution *u*_2_ of our equation. For *t* ≈ 0, it is approximately ∼ *t*, i.e. smaller than ∼ *t*^*c*^ for the dominating solution *u*_2_, and the max *u*_2_ (*t*) occurs significantly earlier.

This maximum is the reason for a well-visible bulge in the middle of the sequence of red dots. Actually, we can see some kind of the bulge for Israel too, but it was short-lived. The coefficient of − *u*_2_ is 1; it is smaller for other considered countries.

Germany: 3/07-5/22. See Figure 14. We begin here with the initial number of total infections 684, which must be subtracted when calculating the *red dots*. This is the moment when the curve began to look stable, i.e. a systematic management began. One has:

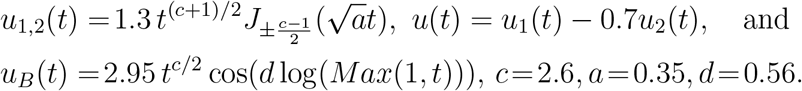

### Japan: 3/20-5/22

See Figure 15. It was a sort of the second wave in Japan, with already 950 total infections on March 20. The curve is discontinuous, but manageable by our *2-phase solution* :

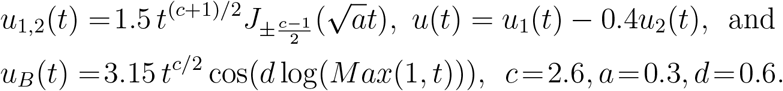

### The Netherlands: 03/13-5/22

Figure 16. The response to *Covid-19* was relatively late in the Netherlands; the number of the total case was 383 on 3/13, the beginning of intensive spread from our perspective.

However, the country perfectly reached the saturation at *t*_*top*_ with a single *u*(*t*) = *u*_1_ (*t*), and then smoothly switched to *phase 2*:

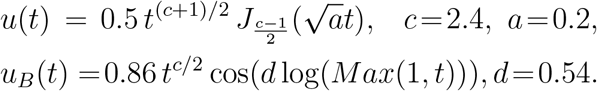

### UK: 03/16-06/13

This country was a challenge for us, though it “eventually” managed to reach phase 2 almost following the “usual pattern”. The *u*-function here is with the same *a, c* as for the Netherlands. Actually the *red dots* match better our *w*-function (from the (*AB*)-mode). However, we prefer to stick to the original *u*(*t*). Then it is combination of two phases *separated by a linear period*, about 10 days. See Figure 17. The parameter *d* = 0.465 is different from that for the Netherlands (0.54). This can be expected; the process toward the saturation of phase 2 was and is slower for UK.

Here *u*(*t*) = *u*_1_ (*t*) is what we used in Figures 3, 10:

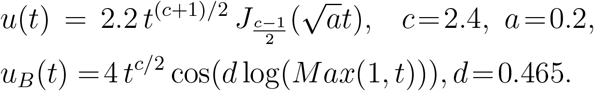

### Second wave in the USA

This solution worked well at least till the middle of September for the second wave in the USA, which began in the middle of June. The accuracy is comparable with what we obtained and discussed in this paper for the first waves in Japan, Israel, Italy, Germany, UK and the Netherlands. Upon subtracting 2.1*M*, the parameters we obtained for the early-middle stage of the 2nd wave in the USA were: *a*_*?*_ = 0.06, *c*_*?*_ = 2.65,

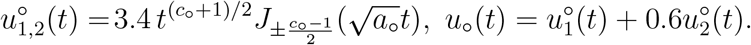

The second phase matched well the following function:

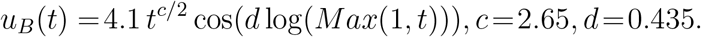

The projected saturation for *u*_*B*_ was given by the formula 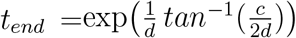. Numerically, *t*_*end*_ = 17.8463, which is 178 days from 06/16: December 11, 2020. See Figure 18. We note that in October, the growth of the number of total cases became linear in the USA.

**Figure 18.**
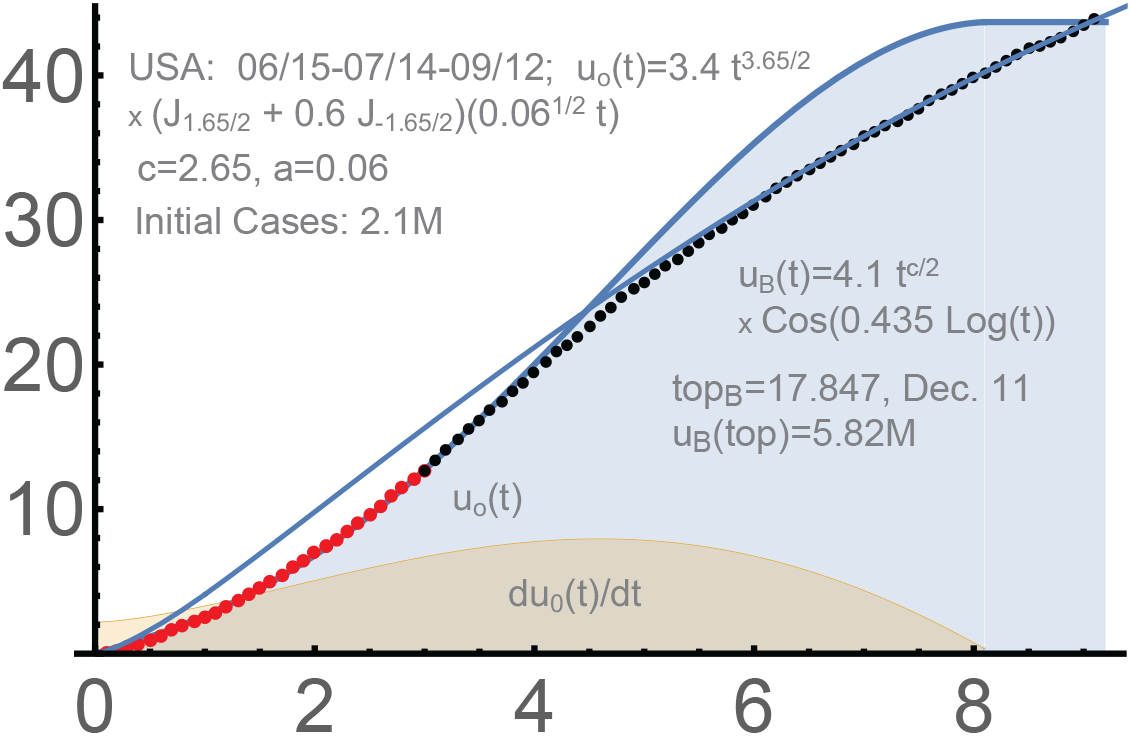
2-phase solution for the 2nd wave in the USA.

To finalize, the best ways to use our curves seem as follows:

- determine *a, c, C* when the spread looks essentially linear;
- update them constantly till the turning point and beyond;
- expect the “bulges” to appear and add the *u*^2^ (*t*) if needed;
- try to adjust the intensity of the measures to match *u*(*t*) ;
- at the turning point, determine *b, D* and the bound *w*(*t*);
- after the saturation at *t*_*top*_, find *d* and switch to *phase 2*,

## 6. Automated forecasting

Generally, our *two-phase solution* serve the best as a forecasting tool if the measures and data are as uniform and “stable” as possible. Then underreporting the number of infections, focusing on symptomatic cases, and inevitable fluctuations with the data may not influence too much the applicability of the *u, w*– curves and *u*_*B*_.

When the situation in the country is fluid, we are supposed to use the programs for constant producing the projections; this appeared the best for the USA. Our software automatically finds, the corresponding curve, determines the current phase, and provides the forecast. The latter can be “linear” if the country/area is in phase 1. For the USA, all 50 states were treated independently, and their *interaction* was subtracted from the total sum of the resulting curves.

The period was taken 03/17-05/27 for all states. We use the data from https://github.com/nytimes/covid-19-data. Our program naturally focuses on the later stages, especially the last 20 days; however, the match with the total number of detected infections appeared perfect almost from 03/17; Figure 19. The *red dots* are the total numbers of detected cases used for finding the parameters, as above.

**Figure 19.**
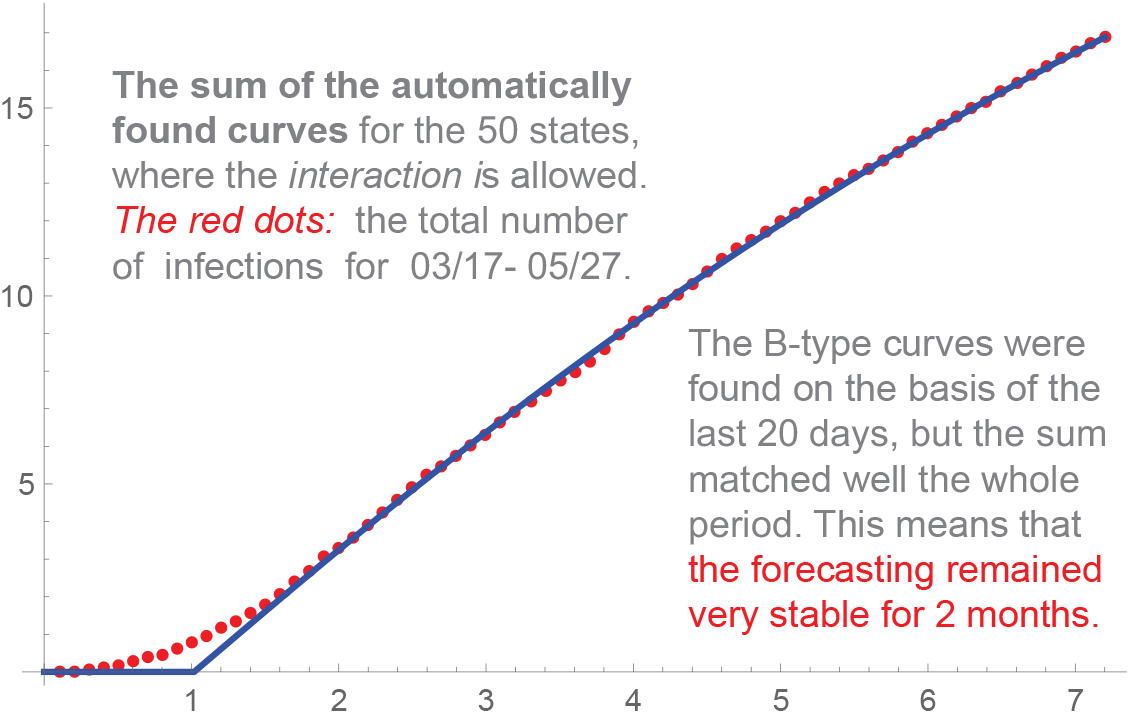
USA, the total of the curves for individual states.

The states that reached the 2nd phase were automatically determined; Figure 20 provides the sum of these curves extended till 07/07 only for these states. The total number of infections is *y* * 100*K*; *x* =days/10. The states are Alaska, Colorado, Connecticut, Delaware, Hawaii, Idaho, Kansas, Kentucky, Louisiana, Massachusetts, Michigan, Missouri, Montana, New Jersey, New York, Oregon, Pennsylvania, Rhode Island, Vermont, Washington, West Virginia, Wyoming as of 05/28. Obviously the situation with these and other states is fluid, and any forecasts can be only conditional. The end of this graph, *x* = 11, corresponds to day 110 from March 17, about July 7.

**Figure 20.**
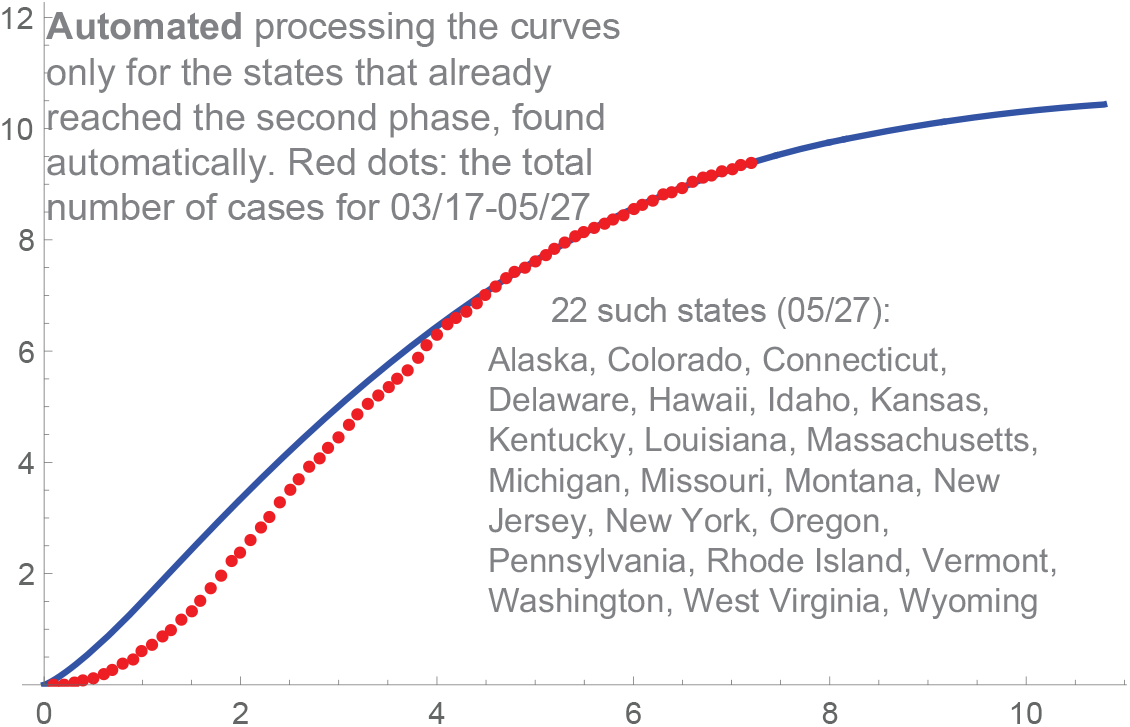
Only for the states reached 2nd stage on 05/27.

The *interaction* is basically allowing our curves in the states reached phase 2 to begin to diminish after their saturation. Currently the projection is that the beginning of October can be when the USA reaches the stage as in Europe in late June or so. Such *iterraction* makes sense, since any states (and countries) benefit from the improvements with the epidemic in their neighbors. We provide one output in Figure 21.

**Figure 21.**
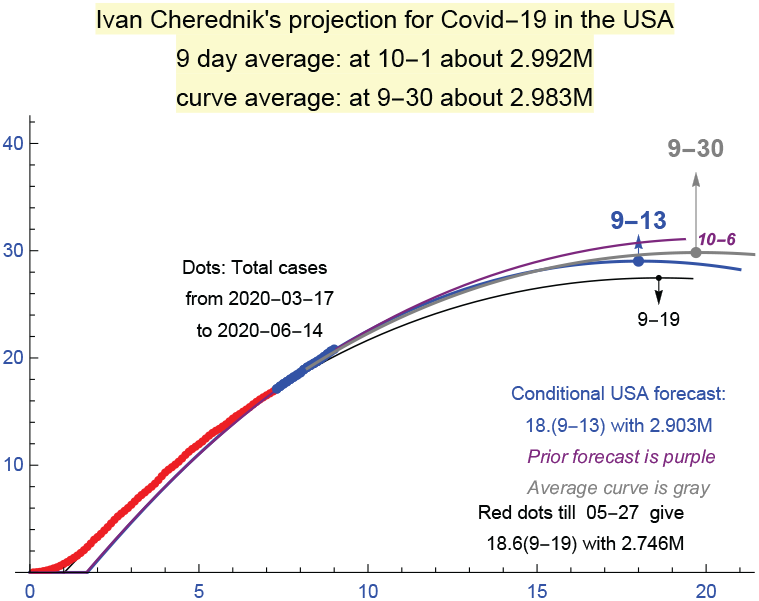
The auto-forecast for the USA as of June 14.

### August-September 2020

There is a variant of this program: the “universal program” for forecasting *late stages* of the waves of *Covid-19* in any countries or groups of countries. It gave quite stable results for Western Europe, but only till the middle of August. Then the second wave began there, presumably due to the end of vacations and the beginning of the school year.

The fact that many schools and businesses were closed in the USA, in contrast to Europe, contributed to a significant reduction of the second wave in the USA, which began around the middle of June. However this trend changed in the middle of September: the number of new daily cases became essentially constant, which corresponds to a linear growth of the total number of cases. In Europe, a strong growth of the total number of (detected) cases began in the end of August, so the program became essentially a linear approximation in September for Europe.

We will provide one of the last outputs, as of 09/22, for the second wave in the USA before the linear growth began there: Fig. 22. The program still found the saturation for the 9-day average of the corresponding curves … but it was 12/28. Here we subtract the initial number of cases, which was 2.1M at 06/16. Actually, only about 15-17 States were in phase 2 as of September 12 and this number dropped to about 11-12 on 9/22.

**Figure 22.**
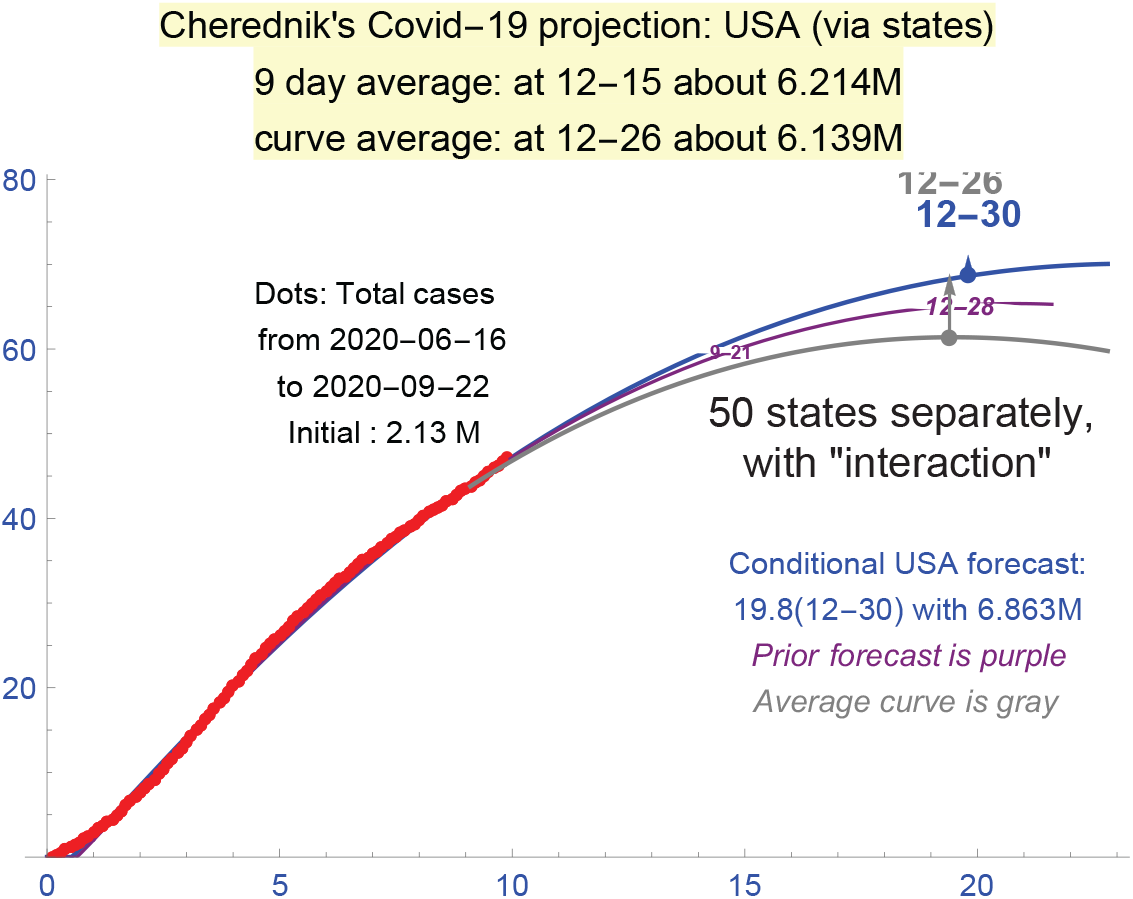
All 50 states separately (9/22).

To conclude, any forecasting required and requires mathematics. Any analysis or discussion of such complex events as epidemics must be based on strict definitions. For instance, comparing different countries and various phases is impossible without solid mathematical methods. This is fully applicable to understanding the efficiency of the measures employed, which can be a difficult task even if the corresponding mathematical tools are adequate.

The management of the spread of *Covid-19* requires constant and sharp monitoring its efficiency, which is possible only if there are reliable mathematical models. Paper [6] was actually the first one focused on the management, especially on the impact of “hard measures”.

Mathematically, our “two-phase solution”, where the role of “hard measures” is assumed the key, provides very accurate modeling the curves of total detected infections. It appeared fully applicable to the second waves, which is quite a confirmation. Without any mathematics, an unusually short break between the first and the second waves in many countries is a clear demonstration that the success with managing the first waves was mostly due to the “hard measures”.

Qualitatively, the importance of the preventive measures is obvious. The problem is to create their quantitative theory and to implement it practically. Our “two-phase solution” seems a very good candidate.

## Supporting information

2 programs and their description

## Data Availability

Extended data and a complete theory are in arxiv 2004.06021v5 (q-bio); the programs forecasting the total number of detected infections for phase 2 are attached.

https://arxiv.org/abs/2004.06021

## Acknowledgements

I’d like to thank ETH-ITS for outstanding hospitality. My special thanks are to Giovanni Felder, Rahul Pandharipande. I thank very much David Kazhdan for important suggestions, Eric Opdam and Alexei Borodin. Funding: partially supported by NSF grant DMS–1901796 and the Simons Foundation. The attached program, producing “phase-2 forecasts”, was tested for Mathematica 11; the usage of exe-files is optional (checked for Windows 10). Use it at your discretion.

## References

1. H. Hethcote, The mathematics of infectious diseases, SIAM Review, 42:4. (2000), 599–653. 2, 7

2. H. Hethcote, and S. Levin, Periodicity in Epidemiological Models, In: Applied Mathematical Ecology. Biomathematics, 18, 193–211, Springer, Berlin, Heidelberg, S. Levin, T. Hallam, L. Gross (eds), 1989. 2

3. S. Meyer, and L. Held, Power-Law models for infectious disease spread, The Annals of Applied Statistics 8:3 (2014), 1612–1639. 2, 3

4. Ph. Strong, Epidemic psychology: a model, Sociology of Health & Illness 12:3 (1990), 249–259. 2

5. S. Cobey, Modeling infectious disease dynamics, Science, 24 Apr 2020; DOI: 10.1126/science.abb5659. 2

6. I. Cherednik, Momentum managing epidemic spread and Bessel functions, Preprint: arxiv 2004.06021v3 (q-bio), 2020 (Chaos, Solitons & Fractals, 2020). 3, 4, 8, 10, 18, 26

7. I. Cherednik, Artificial intelligence approach to momentum risk-taking, Preprint: arxiv 1911.08448v4 (q-fin), 2019. 3, 6

8. R. Carrasco-Hernandez, and R. Jácome, and Y. López Vidal, and S. Ponce de León, Are RNA viruses candidate agents for the next global pandemic? A review, ILAR Journal, 58:3 (2017), 343–358. 8

9. G.N. Watson, A Treatise on the Theory of Bessel Functions, 2nd Edition, Cambridge University Press, Cambridge, 1944. 9

