## Supplementary material for "A Surprising formula for the spread of Covid-19 Under Aggressive Management": 2 programs and their description: programs.pdf

### Auto-forecasting late stages of Covid-19

**Ivan Cherednik**

Let us discuss the programs designed for forecasting late stages of the waves of Covid-19 in any countries or groups of countries. We compare their outputs for the USA with our *2-phase solution* for the USA, which appeared applicable, at least till the middle of September, when the number of new daily cases touched the level comparable with those before the second wave began.

**A. Forecasting late stages**. The programs are designed to be used only at *later stages* of (the waves of) *Covid-19* in any countries, groups of countries, and regions. We provide a "universal program" (any countries) in FOREU.zip, and its special version in FORUS.zip designed for auto-forecasting the total number of cased in the USA via considering all 50 states and calculating the *superposition* of the corresponding projections. These programs are based on the type $B$ formulas for $u_B(t)$, which were used in our *2-phase solution.* They describe *only* later stages: *phase two* in our terminology.

The "universal program" gave quite stable results for Western Europe, but only till the middle of August. Then the second wave began there, presumably due to the end of vacations and the beginning of the school year. The USA program (all states separately) provided stable projections for the saturation till the middle of June, the beginning of the second wave in the USA. Then a significant reduction of the hard measures began. It appeared applicable again in the beginning of September in the USA, with a path to the saturation of the second wave.

The fact that many schools and businesses were closed in the USA, in contrast to Europe, contributed to this. However the trend changed in the middle of September: the number of new daily cases became essentially constant, which corresponds to a linear growth of the number of total cases. In Europe, a strong growth of the total number of (detected) cases began in the end of August, so the program became essentially a linear approximation in September.

**B. Two outputs**. The "universal program" starting with about 9/20 gave the linear approximation for the total number of infections. We provide the output of 9/23 in Figure 1. The projections are for 3 months from the date if the saturation cannot be found during this period.

The second program, based on the consideration of all 50 states, is presumably more reliable. The first run we provide is as of 9/12. The projection was 5.8M in the first 2 weeks of December on top of the initial 2.1M at 06/16. Actually, only about 17 States were in phase 2, so this was not really reliable. This number dropped to about 11 on 9/22. The program still found the saturation for the 9-day average of the corresponding curves but it was 12/28. The trend was toward the linear growth of the total number of cases.

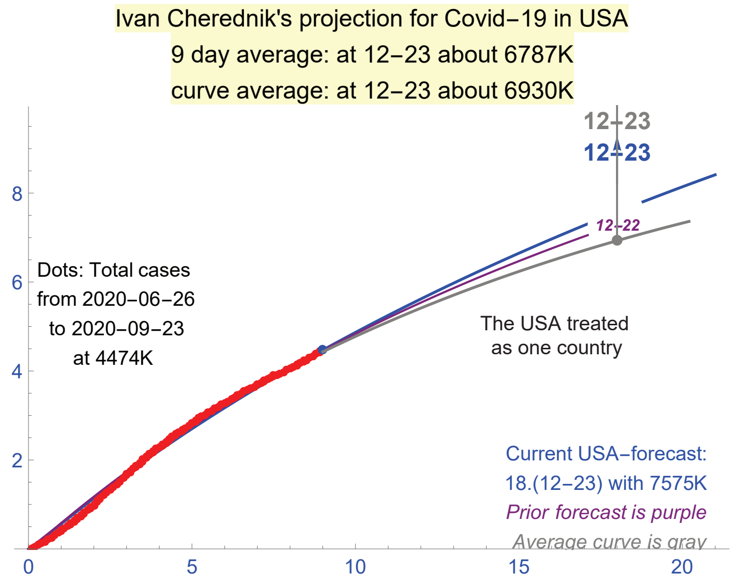


Figure 1. Universal forecast: not via 50 States.

Any reopening of schools and businesses in the USA can and will influence these projections, as occurred in Europe. This already happened in the middle of June, when the USA approached the second phase of the 1st wave, after the hard measures were significantly reduced. Much depends on the virus evolution too. Let us provide two outputs of the program for the USA: Figures 2, 3.

**C. Two phase-solution**. This solution worked well at least till the middle of September for the second wave in the USA. The accuracy is comparable with what we obtained and discussed in this paper for the first waves in Japan, Israel, Italy, Germany, UK and the Netherlands.

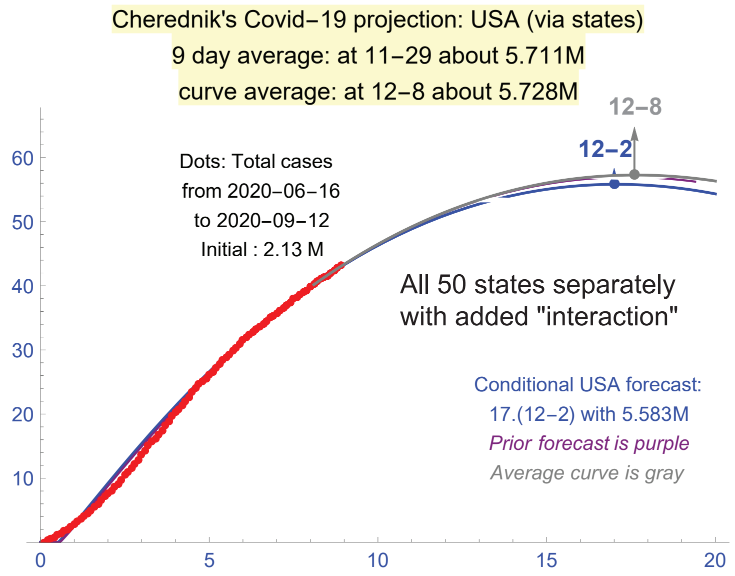


FIGURE 2. All 50 states separately (9/12).

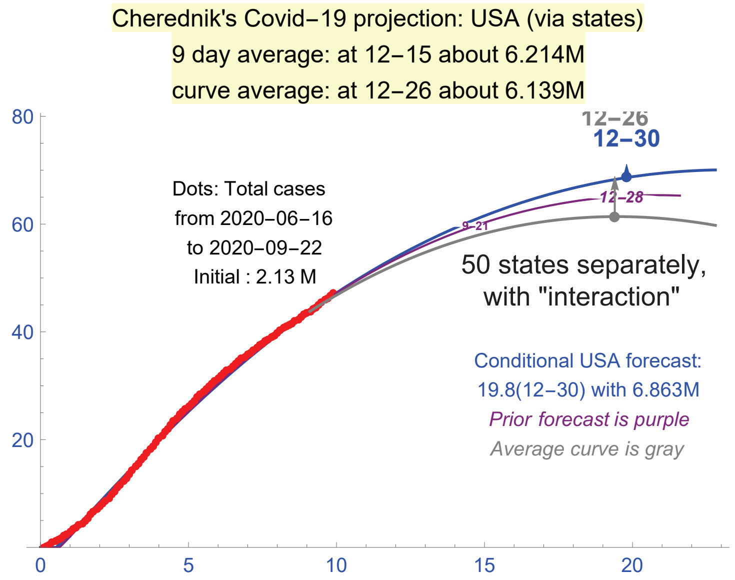


FIGURE 3. All 50 states separately (9/22).

Upon subtracting $2.1M$, the parameters we obtained for the early-middle stage of the 2nd wave in the USA were: $a_\circ = 0.06$, $c_\circ = 2.65$,

$$u^\circ_{1,2}(t) = 3.4\, t^{(c_\circ+1)/2} J_{\pm\frac{c_\circ-1}{2}}(\sqrt{a_\circ}\,t),\ \ u_\circ(t) = u^\circ_1(t) + 0.6 u^\circ_2(t).$$

The second phase matched well the following function:

$$u_B(t) = 4.1\, t^{c/2} \cos(d \log(Max(1,t))), c=2.65, d=0.435.$$

The projected saturation for $u_B$ was given by the formula $t_{end} = \exp\left(\frac{1}{d}\, tan^{-1}(\frac{c}{2d})\right)$. Numerically, $t_{end} = 17.8463$, which is 178 days from 06/16: December 11, 2020. See Figure 4. This date basically matches the auto-projection based on considering all 50 states separately in Figures 2, 3, but the latter are too close to the linear approximation.

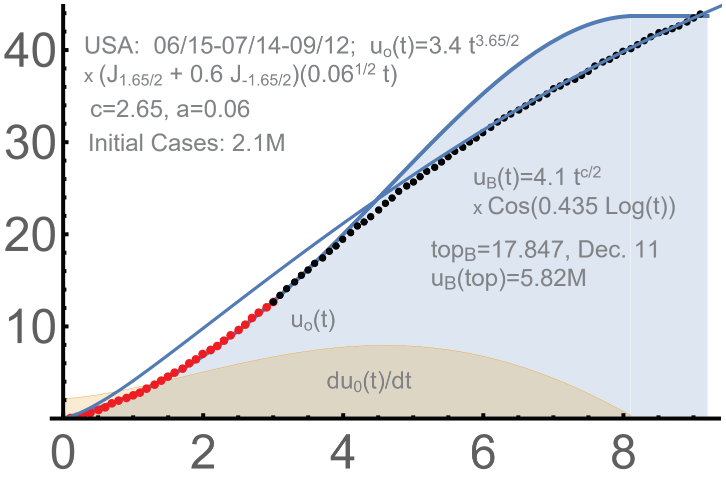


FIGURE 4. 2-phase solution for the 2nd wave in the USA.

**D. Practical matters**. The main programs in *FOREU.zip* are "forwor.txt" and "forworstat.txt". In Mathematica, run "forwor.txt"; it is set for the United States. Upon making $grp = 0$ in "worldfile.txt", it will work for Europe. Some countries were excluded in Europe, mostly due to problems with data. To change this you need to open "forwor.txt" and "forworstat.txt" and find the corresponding places. Mathematica programs are readable.

Generally, the program is "one-click": run the exe-file, after you adjust the path to "math.exe" in your computer in the file "mathpath.txt", and set the countries/regions in "worldfile.txt" following the "README" file.

This is similar for the program for 50 states in the USA. There is no file "worldfile.txt", and currently all states are included. The initial date is set to 06/16/2020. This can be adjusted directly in "forusa.txt" and "forustat.txt": change "datein" (all instances).

The initial date is automatically set to 3 months before "today" in the universal program (for any countries), but not in this one. Here it is the beginning of the wave.

Mathematica 11 is necessary to use these programs. For the universal program, `raw.githubusercontent.com/owid/covid-19-data/` is used: the online access to this site is needed. The names of the countries and regions in "worworld.txt" must be as at this site.
The site `raw.githubusercontent.com/nytimes/covid-19-data/` is used for the program managing the 50 states in the USA.

The *saturation* in these programs is technical; it is supposed to move over time. If the program detects no saturation, then the projections will be provided for a 4 month period; in this case, the program mostly works as a linear extrapolation.

The programs are not for any commercial use; the name of their creator, Ivan Cherednik, and a link to the Journal must be always provided. This is a research tool only. The source file in Mathematica is readable, so you can understand what the programs really do. Please see the README-files.
